# Association of epigenetic age acceleration with MRI biomarkers of aging and Alzheimer’s disease neurodegeneration

**DOI:** 10.64898/2026.01.23.26344650

**Authors:** Linda K. McEvoy, Bowei Zhang, Steve Nguyen, Adam X. Maihofer, Caroline M. Nievergelt, Ramon Casanova, Steve Horvath, Ake T. Lu, Christos Davatzikos, Guray Erus, Susan M. Resnick, Mark A. Espeland, Stephen R. Rapp, Kenneth Beckman, Luigi Ferrucci, Andrea Z. Lacroix, Aladdin H. Shadyab

## Abstract

Epigenetic clocks of biological aging have been associated with cognitive impairment and dementia. Less is known about whether they are associated with an older-appearing brain or with an atrophy pattern associated with dementia. We examined associations of five epigenetic clocks measured at baseline with the Spatial Pattern of Atrophy for Recognition of Brain Aging (SPARE-BA) and the Alzheimer’s Disease Pattern Similarity Score (AD-PS) derived from structural MRIs obtained an average of 8 years later among 1,196 older women. Using linear regression models adjusting for relevant covariates, we observed no associations between any epigenetic clock and accelerated brain aging based on SPARE-BA. We observed a significant association between AgeAccelGrim2 and AD-PS (β = 0.015; 95% CI 0.004 to 0.027; p = 0.01). This association appeared to be primarily driven by the association of a DNA methylation marker of smoking pack years with frontal and temporal lobe volumes. AgeAccelGrim2 was not associated with volumes in regions implicated in early AD (hippocampus and entorhinal cortex). Taken together with prior findings, these results suggest that measures of epigenetic and brain age acceleration capture different aspects of biological aging, and that AgeAccelGrim2 is predictive of neurodegenerative changes associated with smoking that increase risk of dementia.

## INTRODUCTION

Advancing age is associated with cognitive decline and increased risk for Alzheimer’s disease and related dementias (ADRD). Biological aging is heterogeneous, differing between people of the same chronological age and among tissue types within the same individual [1]. Epigenetic clocks have been evaluated as measures of biological aging. First generation epigenetic clocks were developed by assessing DNA methylation (DNAm) patterns predictive of chronological age [2–4]. Second generation clocks assessed DNAm patterns predictive of aging-related health outcomes or mortality [5–7]. A third generation clock developed a DNAm marker to predict 20 year pace of aging across various organ systems [8]. Faster epigenetic aging relative to chronological age, considered a marker of accelerated biological aging, has been referred to as epigenetic age acceleration [4].

Epigenetic age acceleration has been associated with poorer cognitive performance, steeper cognitive decline with age, and increased risk of dementia across several epigenetic clocks, even though these clocks were not trained on cognitive outcomes [9–15]. Less is known about whether epigenetic clocks are associated with structural magnetic resonance imaging (MRI) evidence of advanced brain aging or age-related neurodegenerative disorders such as Alzheimer’s disease (AD).

Aging is accompanied by characteristic changes in brain structure that overlap with, but are distinct from, changes that precede AD dementia [16, 17]. Composite MRI indices have been created to reflect brain signatures of aging and of AD. The Spatial Pattern of Atrophy for Recognition of Brain Aging (SPARE-BA) is an index of brain aging that was derived through application of a high-dimensional pattern classification algorithm to differentiate structural MRI data of younger adults from that of older adults, yielding a predicted brain age score [18, 19]. Older brain-predicted age than chronological age is indicative of accelerated brain aging and is referred to here as SPARE-BA acceleration, or SPARE-BAA.

The AD Pattern Similarity Score (AD-PS), a composite index of the degree to which an individual’s structural MRI reflects that of individuals diagnosed with dementia, was derived by applying high dimensional machine learning to structural MRI measures of gray matter to discriminate scans of healthy older adults from scans of those with AD dementia using data from the Alzheimer’s Disease Neuroimaging Initiative [20]. Higher scores reflect greater similarity with the spatial volumetric MRI pattern observed in participants with AD. The AD-PS strongly predicted future dementia in independent cohorts, including in the Women’s Health Initiative Memory Study (WHIMS) [21, 22]. In WHIMS, the AD-PS showed an area under the curve of 0.89 for discriminating those who developed dementia from cognitively stable women [21, 22].

Greater knowledge of whether epigenetic measures of accelerated aging are associated with structural MRI brain signatures of aging or AD may inform on underlying biological mechanisms and provide more accessible biomarkers for brain changes in aging and AD. To this end, we examined whether epigenetic aging is associated with older brain-predicted age than chronological age or with an atrophy pattern indicative of increased AD risk an average of 8 years later, in a large cohort of community dwelling older women from the WHIMS. We examined five commonly used epigenetic clocks, including two first-generation clocks, Horvath’s intrinsic epigenetic age acceleration (IEAA) and Hannum’s extrinsic epigenetic age acceleration (EEAA), two second-generation clocks, DNAm PhenoAge (AgeAccelPheno) [5] and GrimAge2 (AgeAccelGrim2) [6, 7], and a third-generation clock, Dunedin (P)ace of (A)ging (C)alculated from the (E)pigenome (DunedinPACE) [8].

## RESULTS

Table 1 shows baseline characteristics of the full sample and by AgeAccelGrim2 quartiles. Women who reported never smoking were most likely to be in the lowest AgeAccelGrim2 quartile whereas current smokers were most likely to be in the highest quartile. Women in the lowest quartile of AgeAccelGrim2 were more likely to have higher levels of physical activity and lower body mass index (BMI), whereas those in the highest quartile were more likely to have prevalent diabetes. There were no differences across AgeAccelGrim2 quartiles by hormone treatment (HT) arm, race, ethnicity, education, APOE ε4 carrier status, cardiovascular disease (CVD), or non-melanoma cancer.

**Table 1.** Demographic and clinical characteristics by quartile of AgeAccelGrim2.

| AgeAccelGrim2 | Overall<br>N = 1,196 | Q1<br>[-11.64, -4.35]<br>N = 297 | Q2<br>[-4.35, -2.07]<br>N = 301 | Q3<br>[-2.07, 0.57]<br>N = 298 | Q4<br>[0.57, 19.49]<br>N = 300 | p-value |
| --- | --- | --- | --- | --- | --- | --- |
| **Age, Mean (SD)** | 70 (4) | 69 (3) | 70 (4) | 69 (3) | 70 (4) | 0.003 |
| **Hormone Therapy arm, n (%)** |  |  |  |  |  | 0.2 |
| Estrogen-alone intervention | 228 (19%) | 48 (16%) | 56 (19%) | 71 (24%) | 53 (18%) |  |
| Estrogen-alone placebo | 213 (18%) | 48 (16%) | 53 (18%) | 45 (15%) | 67 (22%) |  |
| Estrogen+Progestin intervention | 365 (31%) | 98 (33%) | 92 (31%) | 87 (29%) | 88 (29%) |  |
| Estrogen+Progestin placebo | 390 (33%) | 103 (35%) | 100 (33%) | 95 (32%) | 92 (31%) |  |
| **Education, n (%)** |  |  |  |  |  | 0.14 |
| Less than high school equivalent | 53 (4.4%) | 11 (3.7%) | 20 (6.6%) | 11 (3.7%) | 11 (3.7%) |  |
| High school diploma or GED | 276 (23%) | 65 (22%) | 63 (21%) | 83 (28%) | 65 (22%) |  |
| Vocational, training school,<br>Some college or associate degree | 469 (39%) | 116 (39%) | 131 (44%) | 106 (36%) | 116 (39%) |  |
| College graduate or higher | 395 (33%) | 104 (35%) | 87 (29%) | 97 (33%) | 107 (36%) |  |
| Missing | 3 | 1 | 0 | 1 | 1 |  |
| **Smoking, n (%)** |  |  |  |  |  | <0.001 |
| Never Smoked | 678 (57%) | 209 (71%) | 202 (67%) | 165 (56%) | 102 (34%) |  |
| Past Smoker | 457 (39%) | 85 (29%) | 96 (32%) | 125 (43%) | 151 (51%) |  |
| Current Smoker | 51 (4.3%) | 0 (0%) | 2 (0.7%) | 4 (1.4%) | 45 (15%) |  |
| Missing | 10 | 3 | 1 | 4 | 2 |  |
| **Smoking, pack years, Mean (SD)** | 8.7 (18.0) | 2.6 (7.5) | 4.9 (14.8) | 7.6 (15.5) | 19.3 (24.9) | <0.001 |
| Missing | 37 | 9 | 11 | 9 | 8 |  |
| **Alcohol consumption, n (%)** |  |  |  |  |  | 0.001* |
| Never | 146 (12.3%) | 38 (12.9%) | 40 (13.4%) | 43 (14.5%) | 25 (8.4%) |  |
| Past | 201 (16.9%) | 27 (9.2%) | 53 (17.7%) | 54 (18.2%) | 67 (22.5%) |  |
| Current light-moderate | 704 (59.3%) | 185 (62.9%) | 174 (58.2%) | 173 (58.2%) | 172 (57.7%) |  |
| Current heavy | 137 (11.5%) | 44 (15.0%) | 32 (10.7%) | 27 (9.1%) | 34 (11.4%) |  |
| Missing | 8 | 3 | 2 | 1 | 2 |  |
| **Race, n (%)** |  |  |  |  |  | 0.4 |
| American Indian/ Alaskan<br>Native | 2 (0.2%) | 0 (0%) | 2 (0.7%) | 0 (0%) | 0 (0%) |  |
| Asian | 21 (1.8%) | 6 (2.0%) | 6 (2.0%) | 4 (1.4%) | 5 (1.7%) |  |
| Black | 55 (4.6%) | 9 (3.0%) | 11 (3.7%) | 14 (4.7%) | 21 (7.0%) |  |
| White | 1,104 (93%) | 279 (94%) | 278 (93%) | 277 (94%) | 270 (91%) |  |
| More than one race | 9 (0.8%) | 3 (1.0%) | 3 (1.0%) | 1 (0.3%) | 2 (0.7%) |  |
| Unknown or not reported | 5 | 0 | 1 | 2 | 2 |  |
| **Ethnicity, n (%)** |  |  |  |  |  | 0.3 |
| not Hispanic or Latino | 1,171 (98%) | 293 (99%) | 296 (98%) | 291 (98%) | 291 (97%) |  |
| Hispanic or Latino | 24 (2.0%) | 3 (1.0%) | 5 (1.7%) | 7 (2.3%) | 9 (3.0%) |  |
| Unknown or not reported | 1 | 1 | 0 | 0 | 0 |  |
| **APOE e4 carrier status, n (%)** |  |  |  |  |  | 0.3 |
| no e4 alleles | 822 (78%) | 204 (74%) | 213 (80%) | 205 (78%) | 200 (80%) |  |
| at least one e4 allele | 230 (22%) | 70 (26%) | 52 (20%) | 57 (22%) | 51 (20%) |  |
| Missing | 144 | 23 | 36 | 36 | 49 |  |
| **Energy expenditure** (recreational physical activity, MET-hours/wk, Mean, SD) | 11 (13) | 13 (12) | 12 (14) | 10 (12) | 10 (13) | <0.001 |
| Missing | 2 | 0 | 0 | 1 | 1 |  |
| **BMI**, Mean (SD) | 28.3 (5.5) | 26.8 (4.9) | 28.1 (5.6) | 29.1 (5.4) | 29.0 (5.6) | <0.001 |
| Missing | 5 | 3 | 1 | 1 | 0 |  |
| **Diabetes, n (%)** | 43 (3.6%) | 4 (1.3%) | 3 (1.0%) | 12 (4.0%) | 24 (8.0%) | <0.001 |
| Missing | 1 | 0 | 1 | 0 | 0 |  |
| **Cardiovascular disease, n (%)** | 32 (2.7%) | 5 (1.7%) | 6 (2.0%) | 8 (2.7%) | 13 (4.3%) | 0.2 |
| **Non-melanoma cancers, n (%)** | 127 (11%) | 38 (13%) | 34 (11%) | 26 (8.7%) | 29 (9.7%) | 0.4 |

Figure 1 shows associations of epigenetic clocks with SPARE-BAA, from fully adjusted models (see Table S1 for results of progressively adjusted models, and Figure S1 for scatter plots). None of the clocks showed significant associations with SPARE-BAA.

**Figure 1.**
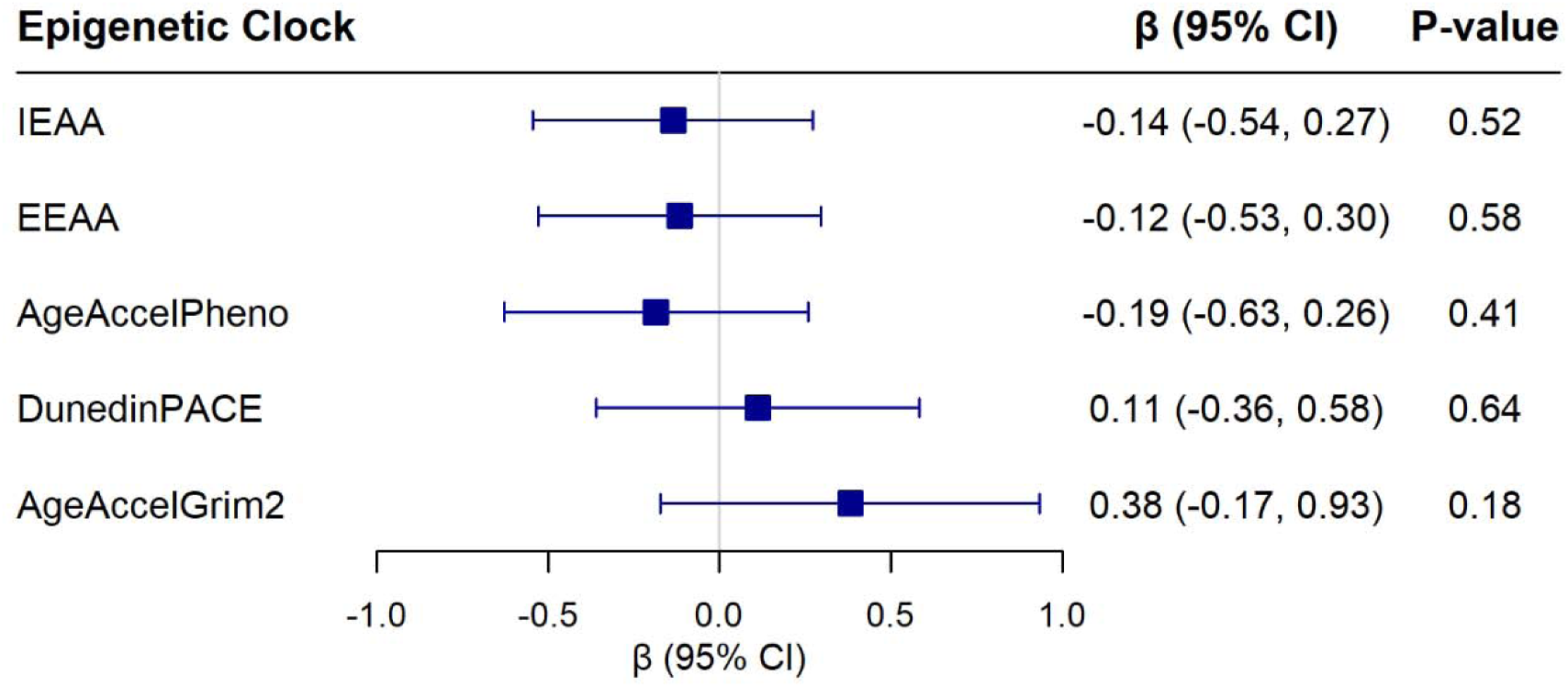
Associations of five epigenetic aging clocks with SPARE-BAA from linear regression models adjusting for chronological age, hormone therapy trial arm, education, smoking status, race, ethnicity, physical activity, BMI, diabetes, cardiovascular disease, cancer, and blood cell composition (models for IEAA and EEAA did not include blood cell composition). Beta coefficients are reported for each 1 standard deviation increase in the epigenetic measure.

Figure 2 shows associations of epigenetic clocks with the AD-PS score, which was natural log-transformed after adding 1 due to its non-normal distribution, from fully adjusted models (see Table S2 for results of progressively adjusted models and Figure S2 for scatter plots). Only AgeAccelGrim2 showed a significant association, which survived correction for multiple comparisons (β = 0.015; 95% CI 0.004 to 0.027; p = 0.01). A one standard deviation increase in AgeAccelGrim2 (4.15) was associated with a 1.5% higher ln(AD-PS+1) score. Of the 10 components of AgeAccelGrim2, only the DNAm marker of smoking pack years was associated with the ln(AD-PS+1) score (β = 0.023; 95% CI 0.011 to 0.035; p < 0.01) (Figure 3). These associations remained significant after further adjustment for smoking pack years based on self-report (AgeAccelGrim2: β = 0.013; 95% CI 0.001 to 0.025; p = 0.04; DNAm smoking pack years: 0.022; 95% CI 0.009 to 0.035; p < 0.01) and after adjustment for alcohol use (AgeAccelGrim2: β = 0.014; 95% CI 0.003 to 0.026; p = 0.01; DNAm smoking pack years: 0.023; 95% CI 0.011 to 0.035; p < 0.01).

**Figure 2.**
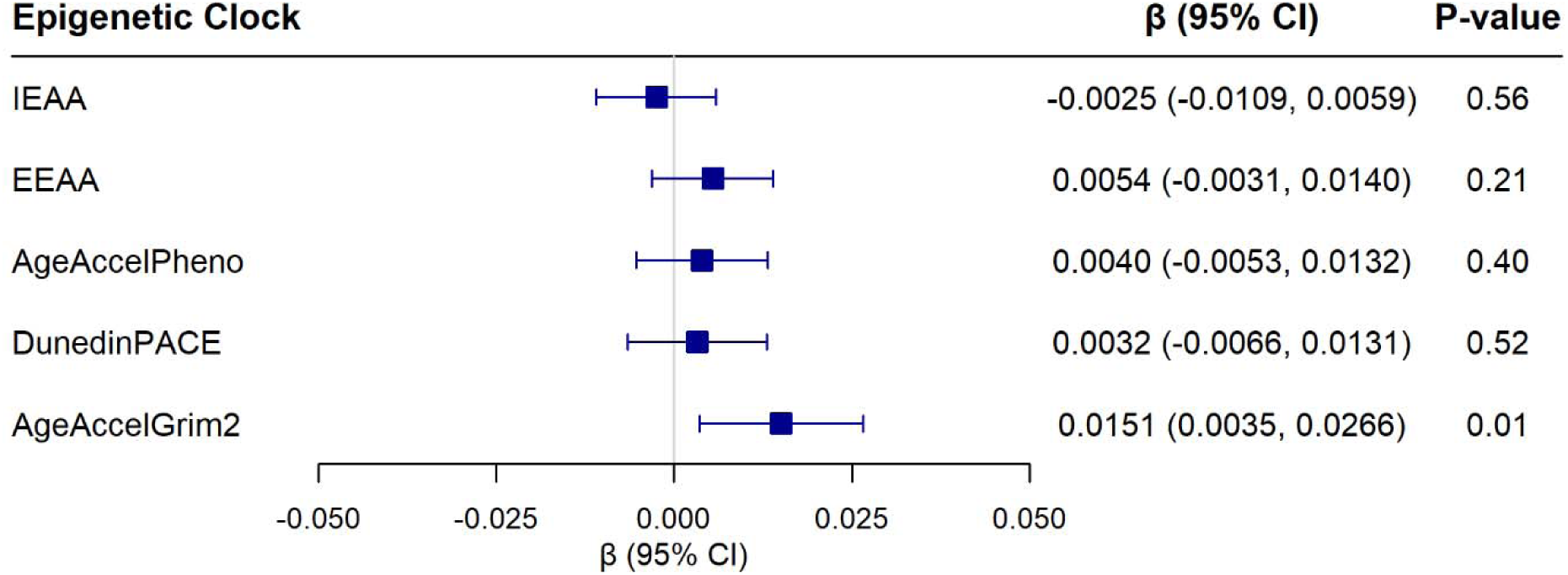
Associations of five epigenetic aging clocks with the Alzheimer’s Disease Pattern Similarity Score [ln (AD-PS + 1)], from linear regression models adjusting for chronological age, hormone therapy trial arm, education, smoking status, race, ethnicity, physical activity, BMI, diabetes, cardiovascular disease, cancer, and blood cell composition (models for IEAA and EEAA did not include blood cell composition). Beta coefficients are reported for each 1 standard deviation increase in the epigenetic measure.

**Figure 3.**
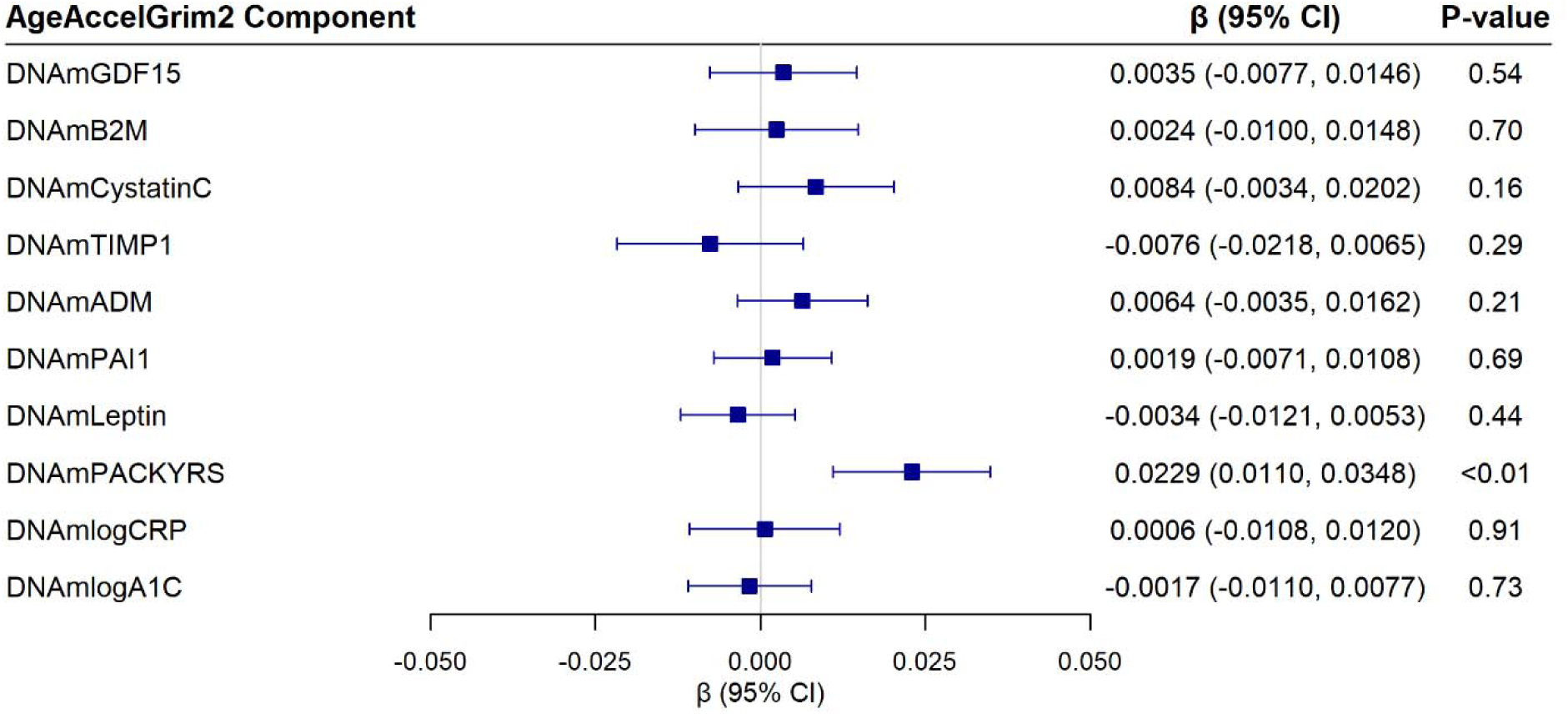
Associations of the ten epigenetic components of the AgeAccelGrim2 clock with the AD-PS score from linear regression models adjusting for chronological age, hormone therapy trial arm, education, smoking status, race, ethnicity, physical activity, BMI, diabetes, cardiovascular disease, cancer, and blood cell composition. Beta coefficients are reported for each 1 standard deviation increase in the epigenetic measure. Abbreviations: DNAm GDF-15: epigenetic marker of growth differentiation factor 15; DNAm B2M: epigenetic marker of beta-2 microglobulin; DNAm Cystatin C: epigenetic marker of cystatin C; DNAm TIMP-1: epigenetic marker of tissue inhibitor metalloproteinase 1; DNAm ADM: epigenetic marker of adrenomedullin; DNAm PAI-1: epigenetic marker of plasminogen activation inhibitor 1; DNAm Leptin: epigenetic marker of leptin; DNAm Packyrs: epigenetic marker of smoking pack-years; DNAm logCRP: epigenetic marker of log-scale high sensitivity C-reactive protein; DNAm logA1C: epigenetic marker of log hemoglobin A1C.

In secondary analyses examining associations of AgeAccelGrim2 with regional brain volumes, AgeAccelGrim2 was inversely associated with total brain volume (β = −2.32; 95% CI −4.47 to −0.17; p = 0.03), frontal lobe volume (β = −1.65; 95% CI −2.81 to −0.49; p<0.01) and temporal lobe volume (β = −0.81; 95% CI −1.49 to −0.13; p = 0.02). There were no associations with other brain regions, including hippocampus and entorhinal cortex (Figure 4). The same pattern of results was observed with DNAm smoking pack-years as the exposure (Figure 5).

**Figure 4.**
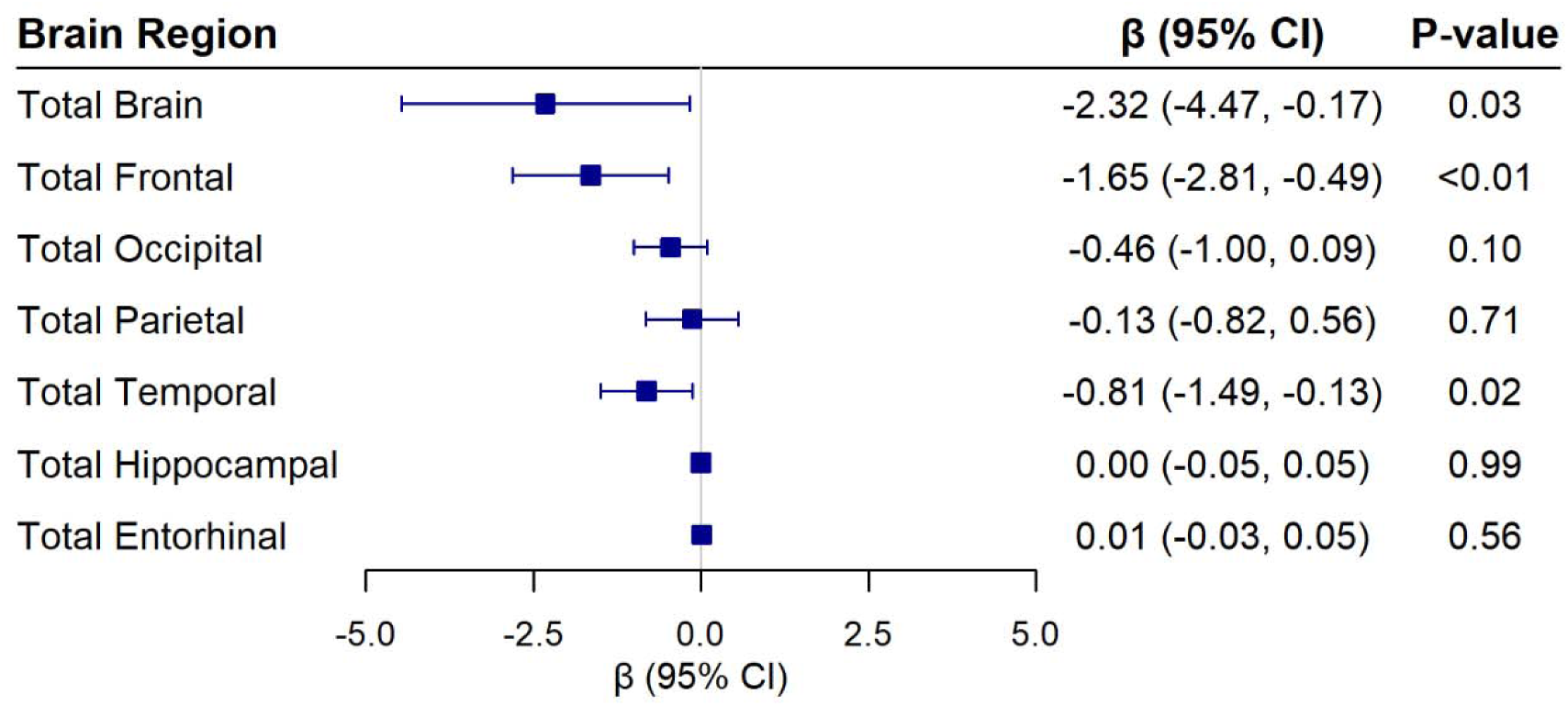
Associations of AgeAccelGrim2 with global and regional brain volumes (in cm^3^) from linear regression models adjusting for chronological age, hormone therapy trial arm, education, smoking status, race, ethnicity, physical activity, BMI, diabetes, cardiovascular disease, cancer, blood cell composition, and intracranial volume. Beta coefficients are reported for each 1 standard deviation increase in the epigenetic measure.

**Figure 5.**
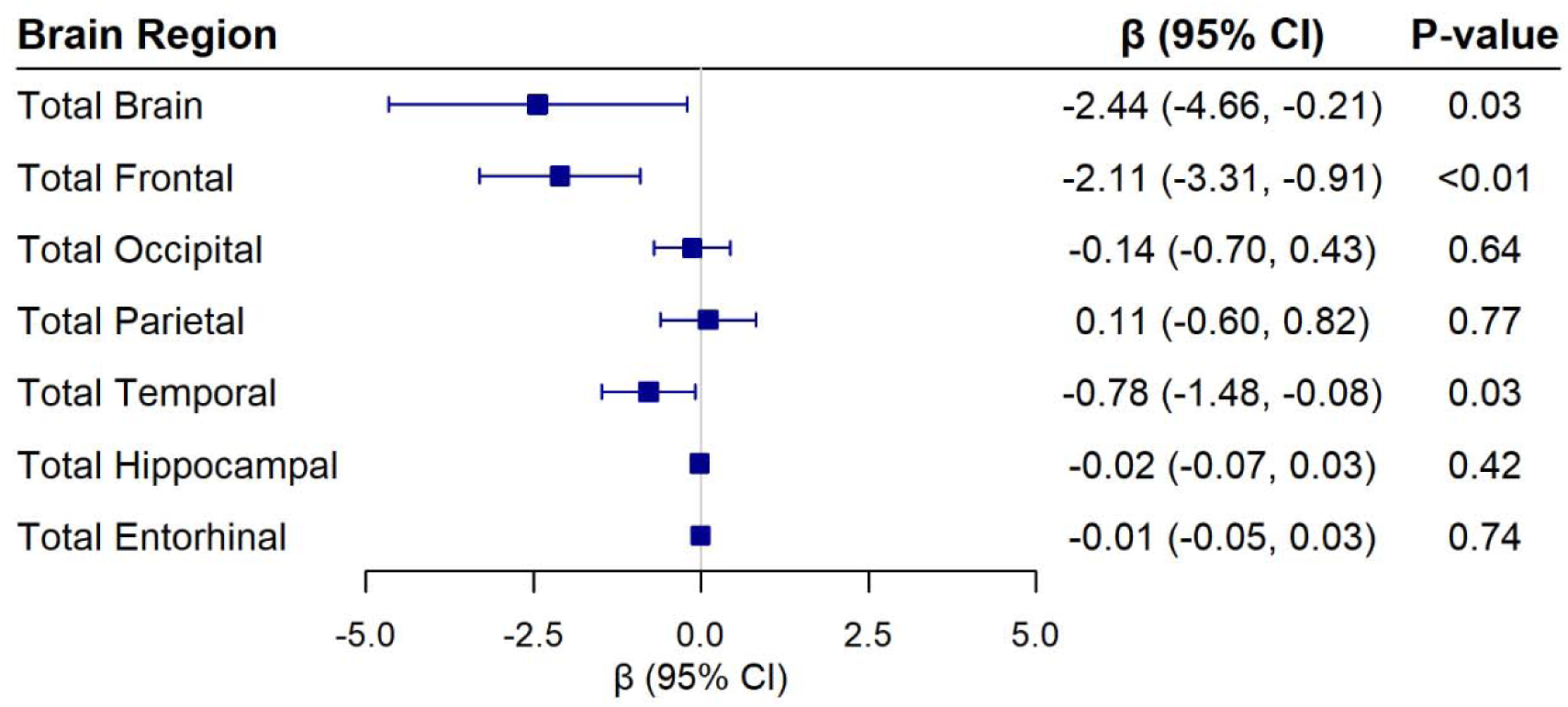
Associations of DNAm SmokingPackYears from AgeAccelGrim2 with global and regional brain volumes (in cm^3^) from linear regression models adjusting for chronological age, hormone therapy trial arm, education, smoking status, race, ethnicity, physical activity, BMI, diabetes, cardiovascular disease, cancer, blood cell composition, and intracranial volume. Beta coefficients are reported for each 1 standard deviation increase in the epigenetic measure.

The exclusion of 46 women with MCI or dementia at time of MRI visit did not change the results (see Table S3; Figure S3). AgeAccelGrim2 continued to show a significant association with the ln(AD-PS+1) score (β = 0.016; 95% CI 0.005 to 0.028; p < 0.01) in these analyses. Associations of AgeAccelGrim2 with the ln(AD-PS+1) and SPARE-BAA did not differ by APOE ε4 status (all *P*-values for interaction > 0.10 in the fully adjusted models; see Tables S4 and S5). Adjustment for time between blood draw and MRI scan did not affect the results (Tables S6 and S7).

## DISCUSSION

Leveraging epigenetic and neuroimaging data from a large, diverse cohort of older women from WHIMS, we examined whether accelerated biological aging as indicated by five well-studied epigenetic clocks was associated with accelerated brain aging or with an AD-pattern of brain atrophy from MRIs obtained 8 years later. We found that none of the epigenetic clocks were associated with accelerated brain aging as measured by the SPARE-BAA. AgeAccelGrim2 was associated with higher AD-PS score, a structural MRI index predictive of dementia. Exploratory analyses of associations of 10 individual components of this clock with the AD-PS score, and with individual brain regions, suggested that the association of AgeAccelGrim2 with the AD-PS score was driven by the association of one component, the epigenetic biomarker of smoking pack years, with reduced frontal and temporal lobe volumes.

Relatively few studies have compared epigenetic clocks with measures of brain age acceleration from structural MRI, and those that have, have generally reported no or weak associations [23]. Across studies, AgeAccelGrim was most likely to correlate with accelerated brain aging [23]. For example, there were no significant associations of IEAA, EEAA, or AgeAccelPheno with SPARE-BAA among 326 middle-aged adults (mean age 50 and 55 years at time of MRI) in the CARDIA study, but AgeAccelGrim was weakly and positively associated with SPARE-BAA measured 10 (r=.16) and 15 (r=.18) years later [24]. Using Cole’s brainAgeR method to predict age from structural MRI data, AgeAccelGrim but not AgeAccelHorvath was positively associated with the brain-predicted age difference (brain-PAD; calculated by subtracting chronological age from predicted brain age) [25, 26]. In contrast, accelerated biological aging was not associated with brain-PAD among 560 men and women aged ≥ 70 years in the ASPREE study for any of the epigenetic clocks examined (Horvath, Hannum, PhenoAge, DunedinPACE, GrimAge, or GrimAge2) [27]. Ferreira et al. reported significant associations of AgeAccelHannum and AgeAccelGrim with brain-PAD among 254 adults with a wide age range (20 – 84 years); AgeAccelHorvath, AgeAccelPheno, or DunedinPACE were not associated with brain-PAD [23]. SPARE-BAA and brain-PAD have been associated with many of the same aging-related health outcomes and mortality as epigenetic clocks [18, 26]. The general lack of correlation of epigenetic and brain age acceleration measures suggests that these metrics capture different aspects of biological aging.

In contrast to the lack of association of any of the epigenetic clocks with SPARE-BAA, we observed a significant association of AgeAccelGrim2 with the AD-PS score: increasing AgeAccelGrim2 was associated with greater similarity to a volumetric pattern predictive of dementia. This is consistent with the results of our prior study among WHIMS women, which showed that, of five epigenetic clocks examined, only AgeAccelGrim2 was significantly associated with incident MCI/dementia after multiple comparison correction [28].

Secondary analyses examining associations of AgeAccelGrim2 with total and regional structural brain measures indicated that AgeAccelGrim2 was associated with lower total brain volume, lower frontal lobe volume, and to a lesser extent, lower temporal lobe volume. AgeAccelGrim2 was not associated with hippocampal or entorhinal volume, brain regions that are strongly implicated in early AD. This may suggest that the association of AgeAccelGrim2 with the AD-PS score may not reflect primarily AD-related pathology.

The GrimAge2 clock predicts biological age based on chronological age, sex, and 10 DNAm-based surrogates of plasma proteins, including an epigenetic signature of smoking pack years. Of these, the DNAm surrogate of smoking pack years showed similar associations as AgeAccelGrim2 with AD-PS and with total brain, frontal and temporal volumes, and similar lack of association with hippocampal and entorhinal cortex volumes.

In the Lothian Birth Cohort, a sample that includes both men and women, AgeAccelGrim2 and DNAm smoking pack years were also associated with total brain volume, as well as frontal and temporal lobe volumes [29]. In that study, effect sizes for the cross-sectional association of AgeAccelGrim2 with brain outcomes were larger than for DNAm smoking packyears, suggesting that other plasma proteins contributed to AgeAccelGrim2 associations with brain outcomes. In our study, associations of DNAm smoking packyears with brain outcomes 8 years later were nominally larger than for AgeAccelGrim2, suggesting that the epigenetic marker of smoking underlies much of the association of AgeAccelGrim2 with AD-PS in older women.

While the number of current smokers in our sample at baseline was very small (less than 5%), there were many former smokers (39%). Epigenetic changes related to smoking have been shown to persist for 30 years after smoking cessation [30]. Furthermore, the epigenetic marker of smoking pack years has been shown to be more strongly predictive of mortality than smoking pack years based on self-report and has been shown to be associated with mortality even among non-smokers [6]. This marker may thus reflect epigenetic changes related to exposure to second-hand smoke or to other environmental toxins. Consistent with this, we found that associations remained significant even after adjustment for smoking pack years.

Smoking is a strong cardiovascular risk factor and is associated with increased risk of dementia [31]. Smoking is known to be associated with accelerated brain aging and with reduced brain volume, particularly in the frontal cortex [32–37]. Taken together, the lack of association of AgeAccelGrim2 with hippocampus and entorhinal cortex volumes, and the strong association with frontal cortex volume that appears to be driven by smoking-related epigenetic changes, suggests that AgeAccelGrim2 may be more predictive of vascular contributions to dementia and to AD pathology. In support of this, we recently observed that AgeAccelGrim2 was associated with a non-specific plasma biomarker of neural injury, neurofilament light chain protein (NfL) at baseline, but not with biomarkers of amyloid pathology, phosphorylated tau at threonine-181 (p-tau181) or phosphorylated tau at threonine-217 (ptau 217) at baseline [38].

Our study has several strengths, including a large sample of older women, the prospective design examining baseline epigenetic clocks in relation to brain outcomes ∼8 years later, and the ability to examine five epigenetic clocks and two validated composite structural neuroimaging measures created to predict brain aging and Alzheimer’s dementia risk. We were able to adjust for many potentially confounding sociodemographic, behavioral, and health variables, and were able to explore associations with regional brain measures in addition to the composite scores.

Limitations include the lack of inclusion of men and the limited racial and ethnic diversity of the sample. Therefore, results may not generalize to men or to women of other races/ethnicities. We also examined only a single MRI signature of brain aging, SPARE-BAA; results may differ for other structural brain age measures.

In summary, our study suggests that none of the five most commonly assessed epigenetic clocks are significantly associated with a measure of accelerated brain aging derived from structural MRIs. We found that AgeAccelGrim2 was associated with the AD-PS score, a volumetric pattern reflective of increased risk of dementia. This association appeared to be driven primarily by smoking-related differences in the frontal and temporal lobes and likely does not reflect processes specific to AD. Together with our prior findings of the association of AgeAccelGrim2 with incident MCI/dementia [28], and with a plasma biomarker of neural injury [38], these findings underscore the robustness of AgeAccelGrim2 as a predictor of cognitive impairment and neurodegeneration.

## METHODS

### Study Population

WHIMS is an ancillary study of the Women’s Health Initiative (WHI) hormone therapy trials. WHIMS was designed to investigate the effects of hormone therapy on cognitive outcomes among 7,479 postmenopausal women ages 65-80 years who were cognitively unimpaired at randomization in 1995-1998. Details on the WHIMS design and protocols have been published [39]. Women were recruited across 39 U.S. clinical centers and randomized to either conjugated equine estrogen (CEE) vs. matching placebo or CEE with medroxyprogesterone acetate (CEE+MPA) vs. matching placebo. Annual in person follow-up for cognitive outcomes continued through 2007. In 2008, WHIMS transitioned to annual telephone-administered cognitive assessments in the WHIMS Epidemiology of Cognitive Health Outcomes (WHIMS-ECHO) study, which followed participants for cognitive outcomes through 2021 [40].Among 7,479 WHIMS participants, we excluded 240 with only one WHIMS cognitive assessment, 519 who did not consent to data sharing of their genetic data through dbGaP, and 304 with no baseline DNA or buffy coat available, leaving 6,416 participants whose baseline biospecimens were used for epigenetics measurement. After quality control (see Supplementary methods), epigenetic data were available for 6,069 participants. Of these, 1,196 women had available brain magnetic resonance imaging data.

### Epigenetic Clocks

DNA methylation was measured in whole blood, collected at WHI baseline, at the University of Minnesota Genomics Center using the Illumina EPIC v2 BeadChip (Illumina, Inc., San Diego, CA), which measures DNA methylation at ∼930,000 cytosine-guanine dinucleotide (CpG) sites. Details of the preprocessing and quality control steps for derivation of epigenetic data are reported in the Supplemental Methods.

We examined five commonly used epigenetic clocks, including intrinsic epigenetic age acceleration (IEAA), a version of the Horvath clock that measures cell-intrinsic methylation changes controlling for age-related differences in blood cell composition, and extrinsic epigenetic age acceleration (EEAA) based on the Hannum clock that incorporates age-related differences in white blood cell-type composition. [41] The Horvath and Hannum clocks are first-generation epigenetic clocks that were trained to predict chronological age. We also examined two second-generation clocks that were trained to predict an aging-related clinical phenotype or mortality. These included DNAm PhenoAge (AgeAccelPheno), which was created by regressing DNAm patterns on a composite clinical phenotype that included biomarkers of kidney, liver, metabolic, and immune function, as well as chronological age [5]; and GrimAge, which was created by regressing a composite biomarker of nine DNAm surrogates of health-related plasma proteins, a DNAm-based estimator of smoking pack-years, age, and sex on mortality risk [6, 7]. A second version of this clock, GrimAge2, additionally incorporated DNAm measures of log-scale C-reactive protein for inflammation and log-scale hemoglobin A1C for glucose metabolism [7]. We used GrimAge2 in our analyses. We also included a third-generation clock, DunedinPACE, which was developed to predict longitudinal change in biomarkers of cardiovascular, metabolic, renal, hepatic, immune, dental, and pulmonary systems among middle-aged adults followed for 20 years [8]. Age acceleration from first- and second-generation clocks was measured as the residual from regressing epigenetic age on chronological age. The online Horvath and Clock Foundation DNAm Age Calculator [2] was used to calculate IEAA, EEAA, AgeAccelPheno, and AgeAccelGrim2. DunedinPACE was calculated using R code available at https://github.com/danbelsky/DunedinPACE.

In secondary analyses, we examined the individual DNAm-based components of AgeAccelGrim2: smoking pack-years (DNAm Packyrs), adrenomedullin (DNAm ADM), beta-2 microglobulin (DNAm B2M), cystatin C (DNAm Cystatin C), growth differentiation factor 15 (DNAm GDF-15), leptin (DNAm Leptin), log-scale high sensitivity C-reactive protein (DNAm logCRP), log-scale hemoglobin A1C (DNAm logA1C), plasminogen activation inhibitor 1 (DNAm PAI-1), and tissue inhibitor metalloproteinase 1 (DNAm TIMP-1).[7]

For interpretation purposes, all epigenetic clock variables were standardized with a mean of 0 and SD of 1. Higher values of first- and second-generation epigenetic clocks indicate accelerated biological aging relative to chronological age, whereas lower values indicate slower biological aging. DunedinPACE measures the pace of biological aging. DunedinPACE values >1 indicate faster pace of aging (e.g., a value of 1.10 indicates a pace of aging 10% faster than the norm for adults in midlife), while values <1 indicate slower pace of aging (e.g., a value of 0.90 indicates a pace of aging 10% slower than the norm for adults in midlife).

### Brain MRI Outcomes

The WHIMS-MRI study was designed to compare MRI findings among women assigned to the intervention vs. placebo arms in the WHIMS trial [42, 43]. WHIMS-MRI was conducted in 14 of the 39 clinical centers that participated in WHIMS during 2005-2006. Details of the MRI protocol, and differences between MRI participants and non-participants have been described [42, 43]. The average time from WHIMS baseline (i.e., the time point of epigenetics measurement) to the MRI scan was 7.97 (0.59) years, (range of 6.43 to 10.20 years).

MRI scans were acquired using a standardized scanning protocol developed by the MRI Quality Control Center in the Department of Radiology, University Pennsylvania, described in detail elsewhere.[42, 43]. Briefly, scans were obtained with a 22 cm field of view and a matrix of 256×256 in 1.5T scanners and included oblique axial spin density/T2-weighted spin echo (TR:3200 ms, TE=30/120 ms, slice thickness= 3 mm), fluid-attenuated inversion recovery (FLAIR) T2-weighted spin echo (TR=8000 ms, TI=2000 ms, TE=100 ms, slice thickness=3 mm), and oblique axial three-dimensional T1-weighted gradient echo (flip angle=30 degrees, TR=21 ms, TE=8 ms, slice thickness=1.5 mm) images from the vertex to the skull base parallel to the anterior commissure-posterior commissure (AC-PC) plane.[21]

A fully automated processing pipeline was applied to each participant’s T1-weighted scan, as previously described.[19, 44, 45]. Preprocessing included correction of magnetic field intensity inhomogeneity and skull-stripping. Each T1 scan was segmented into pre-defined anatomical regions of interest (ROIs) using a multi-atlas, multi-warp label-fusion method, MUSE (MULti-atlas region Segmentation utilizing Ensembles). In the MUSE framework, multiple atlases with semi-automatically extracted ground-truth ROI labels are first warped individually to the target image using two different non-linear registration methods. A spatially adaptive weighted voting strategy is then applied to fuse the ensemble into a final segmentation. Each image was segmented into 145 ROIs spanning the whole brain. Using these data, a summary signature of brain aging, the SPARE-BA index, was derived through application of a high-dimensional pattern classification algorithm to differentiate structural MRI data of younger adults from that of older adults, as previously described[18, 19]. We regressed the SPARE-BA score on chronological age at time of the MRI scan to derive a measure of brain-predicted age difference, in which higher brain-predicted age than chronological age is indicative of accelerated brain aging, which has been referred to as SPARE-BA acceleration, or SPARE-BAA [24].

AD-PS scores were derived by applying high-dimensional machine learning to structural MRI measures of gray matter to discriminate AD scans from those of healthy controls, as previously described [20, 21]. Higher scores reflect greater similarity of an individual’s MRI volumetric pattern to that found in individuals with AD. Given its non-normal distribution, the AD-PS score was natural log-transformed prior to analysis, after adding 1 to avoid values of zero.

Our two primary outcomes were the SPARE-BAA and ln(AD-PS score +1). We examined total and regional brain volumes (cm^3^) including frontal, temporal, parietal, and occipital volumes, as well as hippocampus and entorhinal cortex, as secondary outcomes.

### Covariates

Baseline questionnaires assessed age, race (American Indian/Alaskan Native, Asian, Native Hawaiian or other Pacific Islander, Black, White, more than once race, or unknown/not reported), ethnicity (Hispanic/Latino, not Hispanic/Latino, or unknown/not reported), education (less than high school equivalent, high school diploma or GED, some college, college graduate), smoking status (never smoked, past smoker, current smoker), alcohol use (never consumed alcohol, past use, current light use with up to 7 drinks/week, and heavier use with more than 7 drinks per week); treated diabetes, cardiovascular disease, cancer, and total energy expenditure from recreational physical activity (in MET-hours/week). Height and weight were measured with a stadiometer and balance beam scale, respectively, to calculate BMI. *APOE* ε4 carrier status, defined as presence of at least 1 ε4 allele, was determined in women with available genome-wide genotyping data based on 2 single nucleotide polymorphisms (SNPs), rs429358 and rs7412. These data were available for White women only (n= 1049). Imputation was performed using the 1000 Genomics Project reference panel and the MaCH algorithm implemented in Minimac.[46] Both SNPs had high imputation quality (R^2^ >0.97 for rs429358 and R^2^>0.97 for rs7412).[15] Hormone therapy treatment arm (estrogen alone, estrogen placebo, estrogen plus progestin, or estrogen plus progestin placebo) was also included as a covariate. White blood cell (WBC) counts were estimated for CD4+T cells, CD8+ T cells, natural killer cells, B cells, monocytes, and neutrophils using IDOL [47]. For examination of secondary MRI outcomes of total and regional volumes, we additionally adjusted for total intracranial volume.

### Statistical Analysis

Because we previously found that AgeAccelGrim2 was more strongly associated with MCI/dementia in WHIMS than any of the other epigenetic clocks [28], we examined baseline characteristics by quartiles of AgeAccelGrim2. Differences among quartiles were tested using the Kruskal-Wallis rank sum test for continuous variables, Pearson’s chi-square test of independence for categorical variables, and Fisher’s exact test with simulated p-value (based on 2000 replicates) for categorical variables with low expected counts (e.g. race, ethnicity).

To examine the associations of epigenetic clocks with the primary outcomes, and with total or regional volumes in secondary analyses, we conducted multivariable linear regression models to generate beta estimates and their 95% CIs for 1 SD differences in the epigenetic clock measure. Separate models were fit for each epigenetic clock and each MRI outcome. We describe progressively adjusted models. The minimally adjusted models controlled for chronological age (model 1) as well as race (White, Black and other) and Hispanic ethnicity (yes/no) (model 2). The next model additionally adjusted for hormone therapy trial arm, education, and smoking status (model 3). The full model additionally adjusted for physical activity, BMI, diabetes, cardiovascular disease, cancer, and blood cell composition. Models for IEAA and EEAA did not control for blood cell composition because IEAA was developed to be independent of blood cell composition and EEAA tracks age-related changes in blood cell composition [41]. In examination of global and regional brain volumes in secondary analyses, intracranial volume was also included as a covariate. We also examined individual components of AgeAccelGrim2 in secondary analyses.

There was very little missing data for covariates used in the main analyses (<1%; see Table 1). Missing covariate data were imputed using multivariate imputation by chained equations using the R *mice* package, specifying all study variables with 20 imputations and 20 iterations.

In sensitivity analyses, we further adjusted for smoking pack years and for alcohol use. We also examined associations after excluding 46 women who were diagnosed with MCI or probable dementia prior to the MRI scan. We examined effect modification by *APOE* ε4 carrier status by examining interaction terms of epigenetic clocks and *APOE* ε4 in the models; the significance of the interactions was examined using likelihood ratio tests. Finally, we reran the models for the primary outcomes SPARE-BAA and AD-PS including time between blood draw and MRI scan as a covariate in all progressively adjusted models.

To account for multiple testing across five clocks, we applied a Bonferroni-corrected threshold of *P≤*0.01 (0.05/5) for statistical significance for each primary outcome (SPARE-BAA and ln(AD-PS+1)). We did not consider multiple correction for p-values in secondary analyses, as these analyses were considered exploratory. All analyses were conducted using R statistical software version 4.4.2.

## Data Availability

All data produced in the present study are available upon reasonable request to the authors

## Abbreviations

AD: Alzheimer’s disease (AD)
AD-PS: Alzheimer’s Disease Pattern Similarity Score
AgeAccelGrim2, AgeAccelHannum, AgeAccelHorvath, AgeAccelPheno: age acceleration (older epigenetic than chronological age) based on the GrimAge, Hannum, Horvath and PhenoAge epigenetic clocks, respectively
APOE: Apolipoprotein E
BMI: Body Mass Index
CVD: Cardiovascular disease
DNAm: DNA methylation
DunedinPACE: Dunedin (P)ace of (A)ging (C)alculated from the (E)pigenome
EEAA: extrinsic epigenetic age acceleration
IEAA: intrinsic epigenetic age acceleration
MRI: magnetic resonance imaging
SPARE-BA: Spatial Pattern of Atrophy for Recognition of Brain Aging
SPARE-BAA: Spatial Pattern of Atrophy for Recognition of Brain Aging Acceleration (i.e. older brain-predicted age than chronological age)
WHIMS: Women’s Health Initiative Memory Study

## Author Contributions

L.K.M. and A.H.S. planned and designed the study. A.H.S. obtained funding. L.K.M. performed literature review and drafted the manuscript. BZ conducted the statistical analysis and created the figures and tables. S.N. prepared the dataset and calculated the epigenetic clock data on participants; A.X.M and C.N. led the processing of the epigenetic data for analysis; RC created the AD-PS score. C.D. and G.U. led the processing of brain imaging data and created the SPARE-BA index score. All authors contributed to data interpretation, manuscript review, and editing.

## Acknowledgements

We thank the WHI participants, staff, and investigators. The short list of WHI investigators can be found at: https://www-whi-org.s3.us-west-2.amazonaws.com/wp-content/uploads/WHI-Investigator-Short-List.pdf. The full list of WHI Investigators can be found at the following site: http://www.whi.org/researchers/Documents%2520%2520Write%2520a%2520Paper/WHI%2520Investigator%2520Long%2520List.pdf

## Conflicts of Interest

The authors declare no conflicts of interest related to this study.

## Ethical Statement

Ethical approval for this study was obtained from the University of California San Diego Institutional Review Board.

## Consent

All participants provided written informed consent.

## Funding

This study was funded by grant R01AG074345 from the National Institute on Aging, National Institutes of Health. This study was also supported by funds from a program made possible by residual class settlement funds in the matter of April Krueger v. Wyeth, Inc., Case No. 03-cv-2496 (US District Court, SD of Calif.). S. Nguyen was also supported by the National Institute on Aging (P01 AG052352, 1K99AG082863-02). A. Maihofer was also supported by the Department of Veteran Affairs award #IK2BX006536-02. The WHI Program is funded by the National Heart, Lung, and Blood Institute, National Institutes of Health, and U.S Department of Health and Human Services (75N92021D00001, 75N92021D00002, 75N92021D00003, 75N92021D00004, and 75N92021D00005). The National Heart, Lung, and Blood Institute has representation on the Women’s Health Initiative Steering Committee, which governed the design and conduct of the study, the interpretation of the data, and preparation and approval of manuscripts. The findings and conclusions presented in this paper are those of the author(s) and do not necessarily reflect the views of the NIH or the U.S. Department of Health and Human Services.

## Notes

### Competing Interest Statement

The authors have declared no competing interest.

### Clinical Trial

NCT00685009 (Women's Health Initiative Memory Study)

### Author Declarations

The Institutional Review Board of the University of California San Diego gave ethical approval for this study

## REFERENCES

1. Prattichizzo F, Frige C, Pellegrini V, Scisciola L, Santoro A, Monti D, Rippo MR, Ivanchenko M, Olivieri F and Franceschi C. Organ-specific biological clocks: Ageotyping for personalized anti-aging medicine. Ageing Res Rev. 2024; 96:102253.

2. Horvath S. DNA methylation age of human tissues and cell types. Genome Biol. 2013; 14(10):R115.

3. Hannum G, Guinney J, Zhao L, Zhang L, Hughes G, Sadda S, Klotzle B, Bibikova M, Fan JB, Gao Y, Deconde R, Chen M, Rajapakse I, et al. Genome-wide methylation profiles reveal quantitative views of human aging rates. Mol Cell. 2013; 49(2):359–367.

4. Horvath S and Raj K. DNA methylation-based biomarkers and the epigenetic clock theory of ageing. Nat Rev Genet. 2018; 19(6):371–384.

5. Levine ME, Lu AT, Quach A, Chen BH, Assimes TL, Bandinelli S, Hou L, Baccarelli AA, Stewart JD, Li Y, Whitsel EA, Wilson JG, Reiner AP, et al. An epigenetic biomarker of aging for lifespan and healthspan. Aging (Albany NY). 2018; 10(4):573–591.

6. Lu AT, Quach A, Wilson JG, Reiner AP, Aviv A, Raj K, Hou L, Baccarelli AA, Li Y, Stewart JD, Whitsel EA, Assimes TL, Ferrucci L and Horvath S. DNA methylation GrimAge strongly predicts lifespan and healthspan. Aging (Albany NY). 2019; 11(2):303–327.

7. Lu AT, Binder AM, Zhang J, Yan Q, Reiner AP, Cox SR, Corley J, Harris SE, Kuo PL, Moore AZ, Bandinelli S, Stewart JD, Wang C, et al. DNA methylation GrimAge version 2. Aging (Albany NY). 2022; 14(23):9484–9549.

8. Belsky DW, Caspi A, Corcoran DL, Sugden K, Poulton R, Arseneault L, Baccarelli A, Chamarti K, Gao X, Hannon E, Harrington HL, Houts R, Kothari M, et al. DunedinPACE, a DNA methylation biomarker of the pace of aging. Elife. 2022; 11.

9. Beydoun MA, Shaked D, Tajuddin SM, Weiss J, Evans MK and Zonderman AB. Accelerated epigenetic age and cognitive decline among urban-dwelling adults. Neurology. 2020; 94(6):e613–e625.

10. Bressler J, Marioni RE, Walker RM, Xia R, Gottesman RF, Windham BG, Grove ML, Guan W, Pankow JS, Evans KL, McIntosh AM, Deary IJ, Mosley TH, et al. Epigenetic Age Acceleration and Cognitive Function in African American Adults in Midlife: The Atherosclerosis Risk in Communities Study. J Gerontol A Biol Sci Med Sci. 2020; 75(3):473–480.

11. Maddock J, Castillo-Fernandez J, Wong A, Cooper R, Richards M, Ong KK, Ploubidis GB, Goodman A, Kuh D, Bell JT and Hardy R. DNA Methylation Age and Physical and Cognitive Aging. J Gerontol A Biol Sci Med Sci. 2020; 75(3):504–511.

12. Sugden K, Caspi A, Elliott ML, Bourassa KJ, Chamarti K, Corcoran DL, Hariri AR, Houts RM, Kothari M, Kritchevsky S, Kuchel GA, Mill JS, Williams BS, et al. Association of Pace of Aging Measured by Blood-Based DNA Methylation With Age-Related Cognitive Impairment and Dementia. Neurology. 2022; 99(13):e1402–e1413.

13. Shadyab AH, McEvoy LK, Horvath S, Whitsel EA, Rapp SR, Espeland MA, Resnick SM, Manson JE, Chen JC, Chen BH, Li W, Hayden KM, Bao W, et al. Association of Epigenetic Age Acceleration With Incident Mild Cognitive Impairment and Dementia Among Older Women. J Gerontol A Biol Sci Med Sci. 2022; 77(6):1239–1244.

14. Savin MJ, Wang H, Pei H, Aiello AE, Assuras S, Caspi A, Moffitt TE, Muenning PA, Ryan CP, Shi B, Stern Y, Sugden K, Valeri L and Belsky DW. Association of a pace of aging epigenetic clock with rate of cognitive decline in the Framingham Heart Study Offspring Cohort. Alzheimers Dement (Amst). 2024; 16(4):e70038.

15. Nguyen S, McEvoy LK, Espeland MA, Whitsel EA, Lu A, Horvath S, Manson JE, Rapp SR and Shadyab AH. Associations of epigenetic age estimators with cognitive function trajectories in the Women’s Health Initiative Memory Study. Neurology. 2024; 103(1):e209534.

16. Fjell AM, McEvoy L, Holland D, Dale AM, Walhovd KB and Alzheimer’s Disease Neuroimaging Initiative. Brain changes in older adults at very low risk for Alzheimer’s disease. J Neurosci. 2013; 33(19):8237–8242.

17. Fjell AM, McEvoy L, Holland D, Dale AM, Walhovd KB and Alzheimer’s Disease Neuroimaging Initiative. What is normal in normal aging? Effects of aging, amyloid and Alzheimer’s disease on the cerebral cortex and the hippocampus. Prog Neurobiol. 2014; 117:20–40.

18. Habes M, Janowitz D, Erus G, Toledo JB, Resnick SM, Doshi J, Van der Auwera S, Wittfeld K, Hegenscheid K, Hosten N, Biffar R, Homuth G, Volzke H, et al. Advanced brain aging: relationship with epidemiologic and genetic risk factors, and overlap with Alzheimer disease atrophy patterns. Transl Psychiatry. 2016; 6(4):e775.

19. Habes M, Pomponio R, Shou H, Doshi J, Mamourian E, Erus G, Nasrallah I, Launer LJ, Rashid T, Bilgel M, Fan Y, Toledo JB, Yaffe K, et al. The Brain Chart of Aging: Machine-learning analytics reveals links between brain aging, white matter disease, amyloid burden, and cognition in the iSTAGING consortium of 10,216 harmonized MR scans. Alzheimers Dement. 2021; 17(1):89–102.

20. Casanova R, Hsu FC, Sink KM, Rapp SR, Williamson JD, Resnick SM, Espeland MA and Alzheimer’s Disease Neuroimaging Initiative. Alzheimer’s disease risk assessment using large-scale machine learning methods. PLoS One. 2013; 8(11):e77949.

21. Casanova R, Barnard RT, Gaussoin SA, Saldana S, Hayden KM, Manson JE, Wallace RB, Rapp SR, Resnick SM, Espeland MA, Chen JC, WHIMS-MRI Study Group and Alzheimer’s Disease Neuroimaging Initiative. Using high-dimensional machine learning methods to estimate an anatomical risk factor for Alzheimer’s disease across imaging databases. Neuroimage. 2018; 183:401–411.

22. Casanova R, Hsu FC, Barnard RT, Anderson AM, Talluri R, Whitlow CT, Hughes TM, Griswold M, Hayden KM, Gottesman RF, Wagenknecht LE and Alzheimer’s Disease Neuroimaging Initiative. Comparing data-driven and hypothesis-driven MRI-based predictors of cognitive impairment in individuals from the Atherosclerosis Risk in Communities (ARIC) study. Alzheimers Dement. 2022; 18(4):561–571.

23. Sant’Anna Barbosa Ferreira P, van Dongen J, den Braber A, Boomsma DI, de Geus EJC and van ‘t Ent D. Epigenetic age acceleration in peripheral blood correlates with brain-MRI age acceleration. Brain. 2025.

24. Zheng Y, Habes M, Gonzales M, Pomponio R, Nasrallah I, Khan S, Vaughan DE, Davatzikos C, Seshadri S, Launer L, Sorond F, Sedaghat S, Wainwright D, et al. Mid-life epigenetic age, neuroimaging brain age, and cognitive function: coronary artery risk development in young adults (CARDIA) study. Aging (Albany NY). 2022; 14(4):1691–1712.

25. McLachlan KJJ, Cole JH, Harris SE, Marioni RE, Deary IJ and Gale CR. Attitudes to ageing, biomarkers of ageing and mortality: the Lothian Birth Cohort 1936. J Epidemiol Community Health. 2020; 74(4):377–383.

26. Cole JH, Ritchie SJ, Bastin ME, Valdes Hernandez MC, Munoz Maniega S, Royle N, Corley J, Pattie A, Harris SE, Zhang Q, Wray NR, Redmond P, Marioni RE, et al. Brain age predicts mortality. Mol Psychiatry. 2018; 23(5):1385–1392.

27. Phyo AZZ, Fransquet PD, Wrigglesworth J, Woods RL, Espinoza SE and Ryan J. Sex differences in biological aging and the association with clinical measures in older adults. Geroscience. 2024; 46(2):1775–1788.

28. Nguyen S, Lu A, Horvath S, Espeland MA, Rapp SR, Maihofer AX, Nievergelt CM, LaCroix AZ, McEvoy LK, Resnick SM, Beckman K and Shadyab AH. Epigenetic clocks of biological aging and risk of incident mild cognitive impairment and dementia: the Women’s Health Initiative Memory Study. medRxiv. 2025:2025.2009.2029.25336927.

29. Hillary RF, Stevenson AJ, Cox SR, McCartney DL, Harris SE, Seeboth A, Higham J, Sproul D, Taylor AM, Redmond P, Corley J, Pattie A, Hernandez M, et al. An epigenetic predictor of death captures multi-modal measures of brain health. Mol Psychiatry. 2021; 26(8):3806–3816.

30. Joehanes R, Just AC, Marioni RE, Pilling LC, Reynolds LM, Mandaviya PR, Guan W, Xu T, Elks CE, Aslibekyan S, Moreno-Macias H, Smith JA, Brody JA, et al. Epigenetic signatures of cigarette smoking. Circ Cardiovasc Genet. 2016; 9(5):436–447.

31. Livingston G, Huntley J, Liu KY, Costafreda SG, Selbaek G, Alladi S, Ames D, Banerjee S, Burns A, Brayne C, Fox NC, Ferri CP, Gitlin LN, et al. Dementia prevention, intervention, and care: 2024 report of the Lancet standing Commission. Lancet. 2024; 404(10452):572–628.

32. Smith SM, Vidaurre D, Alfaro-Almagro F, Nichols TE and Miller KL. Estimation of brain age delta from brain imaging. Neuroimage. 2019; 200:528–539.

33. Ning K, Zhao L, Matloff W, Sun F and Toga AW. Association of relative brain age with tobacco smoking, alcohol consumption, and genetic variants. Sci Rep. 2020; 10(1):10.

34. Bittner N, Jockwitz C, Franke K, Gaser C, Moebus S, Bayen UJ, Amunts K and Caspers S. When your brain looks older than expected: combined lifestyle risk and BrainAGE. Brain Struct Funct. 2021; 226(3):621–645.

35. Linli Z, Feng J, Zhao W and Guo S. Associations between smoking and accelerated brain ageing. Prog Neuropsychopharmacol Biol Psychiatry. 2022; 113:110471.

36. Whitsel N, Reynolds CA, Buchholz EJ, Pahlen S, Pearce RC, Hatton SN, Elman JA, Gillespie NA, Gustavson DE, Puckett OK, Dale AM, Eyler LT, Fennema-Notestine C, et al. Long-term associations of cigarette smoking in early mid-life with predicted brain aging from mid- to late life. Addiction. 2022; 117(4):1049–1059.

37. Chang Y, Thornton V, Chaloemtoem A, Anokhin AP, Bijsterbosch J, Bogdan R, Hancock DB, Johnson EO and Bierut LJ. Investigating the relationship between smoking behavior and global brain volume. Biol Psychiatry Glob Open Sci. 2024; 4(1):74–82.

38. Zhang B, McEvoy LK, Nguyen S, Espeland MA, Rapp SR, Horvath S, Lu A, LaCroix AZ, Nievergelt CM, Maihofer AX, Resnick SM, Mielke MM, Beckman K, et al. Epigenetic clocks and longitudinal plasma biomarkers of Alzheimer’s disease. medRxiv. 2025.

39. Shumaker SA, Reboussin BA, Espeland MA, Rapp SR, McBee WL, Dailey M, Bowen D, Terrell T and Jones BN. The Women’s Health Initiative Memory Study (WHIMS): a trial of the effect of estrogen therapy in preventing and slowing the progression of dementia. Control Clin Trials. 1998; 19(6):604–621.

40. Espeland MA, Rapp SR, Manson JE, Goveas JS, Shumaker SA, Hayden KM, Weitlauf JC, Gaussoin SA, Baker LD, Padula CB, Hou L, Resnick SM and WHIMSY and WHIMS-ECHO Study Groups. Long-term Effects on Cognitive Trajectories of Postmenopausal Hormone Therapy in Two Age Groups. J Gerontol A Biol Sci Med Sci. 2017; 72(6):838–845.

41. Chen BH, Marioni RE, Colicino E, Peters MJ, Ward-Caviness CK, Tsai PC, Roetker NS, Just AC, Demerath EW, Guan W, Bressler J, Fornage M, Studenski S, et al. DNA methylation-based measures of biological age: meta-analysis predicting time to death. Aging (Albany NY). 2016; 8(9):1844–1865.

42. Resnick SM, Espeland MA, Jaramillo SA, Hirsch C, Stefanick ML, Murray AM, Ockene J and Davatzikos C. Postmenopausal hormone therapy and regional brain volumes: the WHIMS-MRI Study. Neurology. 2009; 72(2):135–142.

43. Coker LH, Hogan PE, Bryan NR, Kuller LH, Margolis KL, Bettermann K, Wallace RB, Lao Z, Freeman R, Stefanick ML and Shumaker SA. Postmenopausal hormone therapy and subclinical cerebrovascular disease: the WHIMS-MRI Study. Neurology. 2009; 72(2):125–134.

44. Pomponio R, Erus G, Habes M, Doshi J, Srinivasan D, Mamourian E, Bashyam V, Nasrallah IM, Satterthwaite TD, Fan Y, Launer LJ, Masters CL, Maruff P, et al. Harmonization of large MRI datasets for the analysis of brain imaging patterns throughout the lifespan. Neuroimage. 2020; 208:116450.

45. Doshi J, Erus G, Ou Y, Resnick SM, Gur RC, Gur RE, Satterthwaite TD, Furth S, Davatzikos C and Alzheimer’s Disease Neuroimaging Initiative. MUSE: MUlti-atlas region Segmentation utilizing Ensembles of registration algorithms and parameters, and locally optimal atlas selection. Neuroimage. 2016; 127:186–195.

46. Howie B, Fuchsberger C, Stephens M, Marchini J and Abecasis GR. Fast and accurate genotype imputation in genome-wide association studies through pre-phasing. Nat Genet. 2012; 44(8):955–959.

47. Salas LA, Zhang Z, Koestler DC, Butler RA, Hansen HM, Molinaro AM, Wiencke JK, Kelsey KT and Christensen BC. Enhanced cell deconvolution of peripheral blood using DNA methylation for high-resolution immune profiling. Nat Commun. 2022; 13(1):761.

