## Supplementary Materials for "Association of epigenetic age acceleration with MRI biomarkers of aging and Alzheimer’s disease neurodegeneration"

**SUPPLEMENTARY METHODS**

**Methylation Quality Control**

Samples were assessed for quality using 17 control metrics from Illumina. Samples were removed if they did not meet Illumina’s recommended thresholds for each control metric (N=14 removed). Sex was determined by clustering samples on the average intensity values of CpG sites on the X and Y chromosomes.^1^ Samples that fell outside of the female cluster or had a mismatch between reported and detected sex were excluded (N=8 removed). Samples with mean bisulfite intensity values < 4,000 were excluded (N=1 removed). Detection p-values were calculated using out-of-band (OOB) probes.^2^ CpG measurements were set to missing if detection p > 0.05 or <= 3 detection beads (M=66,532 probes). CpG sites were removed if >5% of samples were missing data. Based on the remaining CpG sites, samples were excluded if >5% of CpG sites had missing methylation values (N=43 removed). In the remaining samples, OOB background correction, RELIC dye bias correction, and RCP probe type bias correction were applied using Enmix.^3^

**Sample concordance**

Concordance between samples was measured using the SNP fingerprinting probes built into the array. Illumina SNP probes were converted to genotype data. These genotypes were assessed for pairwise IBD in PLINK. If a sample had a 100% match with an unexpected sample, the pair was compared against genotype array data. Under the assumption that array genotypes represented the 'truth' dataset, the sample from the pair that did not match the array genotype was excluded.

**Relatedness**

Kinship was determined from genotype data using KING.^4^ For each pair of participants with estimated 3rd degree or closer relatedness, one was excluded, with the preference to retain cases. If case status matched, one of the pairs was removed at random.

In WHIMS, there were 7 first degree relative (parent-child, siblings) pairs in the entire data and 53 relative pairs were approximately 3^rd^ degree related (first cousins).

**References**

1.	Murat K, Grüning B, Poterlowicz PW, Westgate G, Tobin DJ, Poterlowicz K. Ewastools: Infinium Human Methylation BeadChip pipeline for population epigenetics integrated into Galaxy. *Gigascience*. 2020;9(5):giaa049. doi:10.1093/gigascience/giaa049

2.	Zhou W, Triche TJ, Laird PW, Shen H. SeSAMe: reducing artifactual detection of DNA methylation by Infinium BeadChips in genomic deletions. *Nucleic Acids Res*. 2018;46(20):e123. doi:10.1093/nar/gky691

3.	Xu Z, Niu L, Taylor JA. The ENmix DNA methylation analysis pipeline for Illumina BeadChip and comparisons with seven other preprocessing pipelines. *Clin Epigenetics*. 2021;13(1):216. doi:10.1186/s13148-021-01207-1

4.	Manichaikul A, Mychaleckyj JC, Rich SS, Daly K, Sale M, Chen WM. Robust relationship inference in genome-wide association studies. *Bioinformatics*. 2010;26(22):2867-2873. doi:10.1093/bioinformatics/btq559

**Table S1. Associations of each epigenetic clock with SPARE-BAA in progressively adjusted models**.

| Epigenetic Clock | SD | Model | β (95% CI) | p |
| --- | --- | --- | --- | --- |
| IEAA | 5.05 | 1 | -0.07 (-0.47, 0.34) | 0.75 |
|  |  | 2 | -0.11 (-0.52, 0.29) | 0.59 |
|  |  | 3 | -0.14 (-0.55, 0.27) | 0.50 |
|  |  | 4 | -0.14 (-0.54, 0.27) | 0.52 |
| EEAA | 6.15 | 1 | -0.01 (-0.41, 0.40) | 0.97 |
|  |  | 2 | -0.08 (-0.49, 0.33) | 0.70 |
|  |  | 3 | -0.09 (-0.50, 0.31) | 0.65 |
|  |  | 4 | -0.12 (-0.53, 0.30) | 0.58 |
| AgeAccelPheno | 6.77 | 1 | -0.01 (-0.41, 0.39) | 0.95 |
|  |  | 2 | -0.05 (-0.45, 0.36) | 0.82 |
|  |  | 3 | -0.08 (-0.48, 0.32) | 0.70 |
|  |  | 4 | -0.19 (-0.63, 0.26) | 0.41 |
| DunedinPACE | 0.10 | 1 | 0.17 (-0.24, 0.57) | 0.42 |
|  |  | 2 | 0.26 (-0.15, 0.66) | 0.22 |
|  |  | 3 | 0.19 (-0.23, 0.60) | 0.38 |
|  |  | 4 | 0.11 (-0.36, 0.58) | 0.64 |
| AgeAccelGrim2 | 4.15 | 1 | 0.34 (-0.06, 0.75) | 0.10 |
|  |  | 2 | 0.44 (0.04, 0.85) | 0.03 |
|  |  | 3 | 0.30 (-0.16, 0.76) | 0.20 |
|  |  | 4 | 0.38 (-0.17, 0.93) | 0.18 |

Abbreviations: SD, standard deviation; IEAA, intrinsic epigenetic age acceleration; EEAA, extrinsic epigenetic age acceleration; AgeAccelPheno, phenotypic age acceleration; DunedinPACE, Dunedin Pace of Aging calculated from the Epigenome; AgeAccelGrim2, accelerated DNA methylation GrimAge version 2. Model 1 adjusted for age. Model 2 adjusted for age, race, and ethnicity. Model 3 adjusted for age, race, ethnicity, hormone trial arm, education, and smoking status. Model 4 adjusted for age, race, ethnicity, hormone trial arm, education, smoking status, physical activity, body mass index, diabetes, cardiovascular disease, non-melanoma cancer, and blood cell composition. For IEAA or EEAA, blood cell composition was not included in the fully adjusted model (see text).

**Table S2. Associations of each epigenetic clock with the ln(AD-PS+1) score**

| Epigenetic Clock | SD | Model | β (95% CI) | p |
| --- | --- | --- | --- | --- |
| IEAA | 5.05 | 1 | -0.0017 (-0.0101, 0.0068) | 0.70 |
|  |  | 2 | -0.0015 (-0.0100, 0.0070) | 0.73 |
|  |  | 3 | -0.0024 (-0.0108, 0.0060) | 0.58 |
|  |  | 4 | -0.0025 (-0.0109, 0.0059) | 0.56 |
| EEAA | 6.15 | 1 | 0.0043 (-0.0041, 0.0128) | 0.31 |
|  |  | 2 | 0.0053 (-0.0032, 0.0139) | 0.22 |
|  |  | 3 | 0.0059 (-0.0026, 0.0144) | 0.17 |
|  |  | 4 | 0.0054 (-0.0031, 0.0140) | 0.21 |
| AgeAccelPheno | 6.77 | 1 | 0.0045 (-0.0040, 0.0129) | 0.30 |
|  |  | 2 | 0.0048 (-0.0037, 0.0133) | 0.27 |
|  |  | 3 | 0.0038 (-0.0046, 0.0122) | 0.37 |
|  |  | 4 | 0.0040 (-0.0053, 0.0132) | 0.40 |
| DunedinPACE | 0.10 | 1 | 0.0051 (-0.0034, 0.0135) | 0.24 |
|  |  | 2 | 0.0042 (-0.0044, 0.0127) | 0.34 |
|  |  | 3 | 0.0027 (-0.0059, 0.0113) | 0.53 |
|  |  | 4 | 0.0032 (-0.0066, 0.0131) | 0.52 |
| AgeAccelGrim2 | 4.15 | 1 | 0.0152 (0.0068, 0.0237) | <0.01 |
|  |  | 2 | 0.0144 (0.0059, 0.0229) | <0.01 |
|  |  | 3 | 0.0112 (0.0016, 0.0207) | 0.02 |
|  |  | 4 | 0.0151 (0.0035, 0.0266) | 0.01 |

Abbreviations: SD, standard deviation; IEAA, intrinsic epigenetic age acceleration; EEAA, extrinsic epigenetic age acceleration; AgeAccelPheno, phenotypic age acceleration; DunedinPACE, Dunedin Pace of Aging calculated from the Epigenome; AgeAccelGrim2, accelerated DNA methylation GrimAge version 2. Model 1 adjusted for age. Model 2 adjusted for age, race, and ethnicity. Model 3 adjusted for age, race, ethnicity, hormone trial arm, education, and smoking status. Model 4 adjusted for age, race, ethnicity, hormone trial arm, education, smoking status, physical activity, body mass index, diabetes, cardiovascular disease, non-melanoma cancer, and blood cell composition. For IEAA or EEAA, blood cell composition was not included in the fully adjusted model (see text).

**Table S3. Associations of each epigenetic clock with the ln(AD-PS+1) score after excluding the 46 women with MCI/dementia at the MRI visit**.

| EpiAgeAccel | SD | Model | β (95% CI) | p |
| --- | --- | --- | --- | --- |
| IEAA | 5.04 | 1 | -0.0013 (-0.0098, 0.0072) | 0.76 |
|  |  | 2 | -0.0008 (-0.0093, 0.0078) | 0.86 |
|  |  | 3 | -0.0015 (-0.0099, 0.0069) | 0.73 |
|  |  | 4 | -0.0017 (-0.0101, 0.0068) | 0.70 |
| EEAA | 6.11 | 1 | 0.0033 (-0.0052, 0.0117) | 0.45 |
|  |  | 2 | 0.0045 (-0.0040, 0.0130) | 0.30 |
|  |  | 3 | 0.0053 (-0.0031, 0.0138) | 0.22 |
|  |  | 4 | 0.0050 (-0.0035, 0.0135) | 0.25 |
| AgeAccelPheno | 6.73 | 1 | 0.0049 (-0.0035, 0.0134) | 0.25 |
|  |  | 2 | 0.0055 (-0.0030, 0.0140) | 0.20 |
|  |  | 3 | 0.0048 (-0.0037, 0.0132) | 0.27 |
|  |  | 4 | 0.0054 (-0.0038, 0.0146) | 0.25 |
| DunedinPACE | 0.10 | 1 | 0.0040 (-0.0044, 0.0125) | 0.35 |
|  |  | 2 | 0.0033 (-0.0052, 0.0118) | 0.45 |
|  |  | 3 | 0.0019 (-0.0066, 0.0105) | 0.66 |
|  |  | 4 | 0.0016 (-0.0082, 0.0114) | 0.75 |
| AgeAccelGrim2 | 4.14 | 1 | 0.0158 (0.0074, 0.0243) | <0.01 |
|  |  | 2 | 0.0150 (0.0065, 0.0236) | <0.01 |
|  |  | 3 | 0.0124 (0.0029, 0.0220) | 0.01 |
|  |  | 4 | 0.0163 (0.0048, 0.0277) | <0.01 |

Abbreviations: SD, standard deviation; IEAA, intrinsic epigenetic age acceleration; EEAA, extrinsic epigenetic age acceleration; AgeAccelPheno, phenotypic age acceleration; DunedinPACE, Dunedin Pace of Aging calculated from the Epigenome; AgeAccelGrim2, accelerated DNA methylation GrimAge version 2. Model 1 adjusted for age. Model 2 adjusted for age, race, and ethnicity. Model 3 adjusted for age, race, ethnicity, hormone trial arm, education, and smoking status. Model 4 adjusted for age, race, ethnicity, hormone trial arm, education, smoking status, physical activity, body mass index, diabetes, cardiovascular disease, non-melanoma cancer, and blood cell composition. For IEAA or EEAA, blood cell composition was not included in the fully adjusted model (see text).

**Table S4. Associations of each epigenetic clock with SPARE-BAA stratified by APOE ε4 carrier status.**

|  |  |  | | Carrier | |  | Non-carrier |  |
| --- | --- | --- | --- | --- | --- | --- | --- | --- |
| EpiAgeAccel | | Model | SD | β (95% CI) |  | SD | β (95% CI) | P interaction |
| IEAA | | 1 | 4.71 | 0.60 (-0.34, 1.53) |  | 5.14 | -0.29 (-0.77, 0.19) | 0.09 |
|  | | 2 |  | 0.54 (-0.40, 1.47) |  |  | -0.33 (-0.81, 0.15) | 0.10 |
|  | | 3 |  | 0.56 (-0.39, 1.51) |  |  | -0.31 (-0.79, 0.17) | 0.11 |
| EEAA | | 1 | 6.39 | -0.28 (-1.23, 0.67) |  | 6.02 | 0.07 (-0.41, 0.55) | 0.59 |
|  | | 2 |  | -0.35 (-1.31, 0.60) |  |  | 0.06 (-0.42, 0.54) | 0.52 |
|  | | 3 |  | -0.33 (-1.31, 0.65) |  |  | 0.02 (-0.47, 0.50) | 0.57 |
| AgeAccelPheno | | 1 | 6.62 | -0.23 (-1.17, 0.71) |  | 6.77 | 0.01 (-0.47, 0.49) | 0.72 |
|  | | 2 |  | -0.42 (-1.38, 0.54) |  |  | -0.02 (-0.50, 0.46) | 0.66 |
|  | | 3 |  | 0.00 (-1.05, 1.05) |  |  | -0.26 (-0.79, 0.26) | 0.78 |
| DunedinPACE | | 1 | 0.11 | 0.07 (-0.87, 1.01) |  | 0.10 | 0.38 (-0.10, 0.86) | 0.56 |
|  | | 2 |  | -0.08 (-1.06, 0.90) |  |  | 0.33 (-0.16, 0.82) | 0.56 |
|  | | 3 |  | -0.23 (-1.33, 0.87) |  |  | 0.22 (-0.34, 0.78) | 0.45 |
| AgeAccelGrim2 | | 1 | 3.96 | 0.13 (-0.81, 1.06) |  | 4.05 | 0.58 (0.10, 1.06) | 0.37 |
|  | | 2 |  | -0.37 (-1.48, 0.73) |  |  | 0.52 (-0.02, 1.07) | 0.30 |
|  | | 3 |  | 0.08 (-1.26, 1.42) |  |  | 0.48 (-0.18, 1.13) | 0.30 |

Abbreviations: SD, standard deviation; IEAA, intrinsic epigenetic age acceleration; EEAA, extrinsic epigenetic age acceleration; AgeAccelPheno, phenotypic age acceleration; DunedinPACE, Dunedin Pace of Aging calculated from the Epigenome; AgeAccelGrim2, accelerated DNA methylation GrimAge version 2. Model 1 adjusted for age. Model 2 adjusted for age, race, and ethnicity. Model 3 adjusted for age, race, ethnicity, hormone trial arm, education, and smoking status. Model 4 adjusted for age, race, ethnicity, hormone trial arm, education, smoking status, physical activity, body mass index, diabetes, cardiovascular disease, non-melanoma cancer, and blood cell composition. For IEAA or EEAA, blood cell composition was not included in the fully adjusted model (see text).

P-interactions were obtained from likelihood ratio test comparing the main effects model with a model including the interaction term between scaled epigenetic clock and APOE ε4 carrier status

**Table S5. Associations of each epigenetic clock with the ln(AD-PS+1) score stratified by APOE ε4 carrier status.**

|  |  | Carrier | |  | Non-carrier | |  |
| --- | --- | --- | --- | --- | --- | --- | --- |
| EAA | Model | SD | β (95% CI) |  | SD | β (95% CI) | P interaction |
| IEAA | 1 | 4.71 | 0.0017 (-0.0188, 0.0222) |  | 5.14 | -0.0001 (-0.0102, 0.0099) | 0.87 |
|  | 2 |  | 0.0003 (-0.0204, 0.0209) |  |  | -0.0013 (-0.0113, 0.0086) | 0.81 |
|  | 3 |  | 0.0001 (-0.0209, 0.0211) |  |  | -0.0008 (-0.0108, 0.0091) | 0.88 |
| EEAA | 1 | 6.39 | 0.0162 (-0.0044, 0.0369) |  | 6.02 | 0.0034 (-0.0066, 0.0135) | 0.30 |
|  | 2 |  | 0.0167 (-0.0042, 0.0377) |  |  | 0.0039 (-0.0061, 0.0138) | 0.28 |
|  | 3 |  | 0.0169 (-0.0045, 0.0383) |  |  | 0.0028 (-0.0072, 0.0128) | 0.25 |
| AgeAccelPheno | 1 | 6.62 | 0.0011 (-0.0195, 0.0217) |  | 6.77 | 0.0073 (-0.0028, 0.0173) | 0.57 |
|  | 2 |  | -0.0014 (-0.0225, 0.0197) |  |  | 0.0058 (-0.0042, 0.0157) | 0.69 |
|  | 3 |  | -0.0015 (-0.0255, 0.0224) |  |  | 0.0047 (-0.0061, 0.0155) | 0.79 |
| DunedinPACE | 1 | 0.11 | 0.0015 (-0.0191, 0.0221) |  | 0.10 | 0.0040 (-0.0061, 0.0141) | 0.80 |
|  | 2 |  | -0.0042 (-0.0257, 0.0172) |  |  | 0.0032 (-0.0069, 0.0133) | 0.75 |
|  | 3 |  | -0.0074 (-0.0325, 0.0177) |  |  | 0.0032 (-0.0084, 0.0148) | 0.65 |
| AgeAccelGrim2 | 1 | 3.96 | 0.0196 (-0.0008, 0.0400) |  | 4.05 | 0.0131 (0.0030, 0.0232) | 0.53 |
|  | 2 |  | 0.0111 (-0.0132, 0.0353) |  |  | 0.0099 (-0.0013, 0.0212) | 0.57 |
|  | 3 |  | 0.0112 (-0.0193, 0.0418) |  |  | 0.0129 (-0.0006, 0.0264) | 0.54 |

Abbreviations: SD, standard deviation; IEAA, intrinsic epigenetic age acceleration; EEAA, extrinsic epigenetic age acceleration; AgeAccelPheno, phenotypic age acceleration; DunedinPACE, Dunedin Pace of Aging calculated from the Epigenome; AgeAccelGrim2, accelerated DNA methylation GrimAge version 2. Model 1 adjusted for age. Model 2 adjusted for age, race, and ethnicity. Model 3 adjusted for age, race, ethnicity, hormone trial arm, education, and smoking status. Model 4 adjusted for age, race, ethnicity, hormone trial arm, education, smoking status, physical activity, body mass index, diabetes, cardiovascular disease, non-melanoma cancer, and blood cell composition. For IEAA or EEAA, blood cell composition was not included in the fully adjusted model (see text).

P-interactions were obtained from likelihood ratio test comparing the main effects model with a model including the interaction term between scaled epigenetic clock and APOE ε4 carrier status.

**Table S6. Associations of each epigenetic clock with SPARE-BAA adjusted for time interval since baseline**.

| Epigenetic Clock | SD | Model | β (95% CI) | p |
| --- | --- | --- | --- | --- |
| IEAA | 5.05 | 1 | -0.06 (-0.46, 0.34) | 0.78 |
|  |  | 2 | -0.10 (-0.51, 0.30) | 0.62 |
|  |  | 3 | -0.13 (-0.54, 0.28) | 0.53 |
|  |  | 4 | -0.13 (-0.53, 0.28) | 0.55 |
| EEAA | 6.15 | 1 | -0.03 (-0.44, 0.37) | 0.88 |
|  |  | 2 | -0.10 (-0.51, 0.31) | 0.62 |
|  |  | 3 | -0.11 (-0.52, 0.30) | 0.59 |
|  |  | 4 | -0.14 (-0.55, 0.28) | 0.52 |
| AgeAccelPheno | 6.77 | 1 | -0.04 (-0.44, 0.37) | 0.86 |
|  |  | 2 | -0.07 (-0.47, 0.34) | 0.74 |
|  |  | 3 | -0.10 (-0.51, 0.31) | 0.63 |
|  |  | 4 | -0.19 (-0.63, 0.26) | 0.41 |
| DunedinPACE | 0.10 | 1 | 0.14 (-0.26, 0.55) | 0.48 |
|  |  | 2 | 0.24 (-0.17, 0.65) | 0.26 |
|  |  | 3 | 0.17 (-0.25, 0.58) | 0.43 |
|  |  | 4 | 0.11 (-0.36, 0.58) | 0.65 |
| AgeAccelGrim2 | 4.15 | 1 | 0.32 (-0.09, 0.72) | 0.13 |
|  |  | 2 | 0.42 (0.01, 0.83) | 0.05 |
|  |  | 3 | 0.27 (-0.19, 0.74) | 0.25 |
|  |  | 4 | 0.37 (-0.18, 0.92) | 0.19 |

Abbreviations: SD, standard deviation; IEAA, intrinsic epigenetic age acceleration; EEAA, extrinsic epigenetic age acceleration; AgeAccelPheno, phenotypic age acceleration; DunedinPACE, Dunedin Pace of Aging calculated from the Epigenome; AgeAccelGrim2, accelerated DNA methylation GrimAge version 2. Model 1 adjusted for age and time between blood draw and MRI assessment. Model 2 adjusted for age, time interval between blood draw and MRI assessment, race, and ethnicity. Model 3 adjusted for age, time interval between blood draw and MRI assessment, race, ethnicity, hormone trial arm, education, and smoking status. Model 4 adjusted for age, time interval between blood draw and MRI assessment, race, ethnicity, hormone trial arm, education, smoking status, physical activity, body mass index, diabetes, cardiovascular disease, non-melanoma cancer, and blood cell composition. For IEAA or EEAA, blood cell composition was not included in the fully adjusted model (see text).

**Table S7. Associations of each epigenetic clock with the ln(AD-PS+1) score adjusted for time interval since baseline**.

| Epigenetic Clock | SD | Model | β (95% CI) | p |
| --- | --- | --- | --- | --- |
| IEAA | 5.05 | 1 | -0.0020 (-0.0104, 0.0064) | 0.64 |
|  |  | 2 | -0.0020 (-0.0105, 0.0065) | 0.65 |
|  |  | 3 | -0.0030 (-0.0114, 0.0055) | 0.49 |
|  |  | 4 | -0.0031 (-0.0115, 0.0054) | 0.48 |
| EEAA | 6.15 | 1 | 0.0054 (-0.0030, 0.0139) | 0.21 |
|  |  | 2 | 0.0064 (-0.0021, 0.0150) | 0.14 |
|  |  | 3 | 0.0071 (-0.0014, 0.0155) | 0.10 |
|  |  | 4 | 0.0066 (-0.0019, 0.0151) | 0.13 |
| AgeAccelPheno | 6.77 | 1 | 0.0056 (-0.0028, 0.0141) | 0.19 |
|  |  | 2 | 0.0059 (-0.0026, 0.0144) | 0.17 |
|  |  | 3 | 0.0050 (-0.0034, 0.0134) | 0.24 |
|  |  | 4 | 0.0040 (-0.0052, 0.0132) | 0.39 |
| DunedinPACE | 0.10 | 1 | 0.0061 (-0.0023, 0.0146) | 0.16 |
|  |  | 2 | 0.0053 (-0.0033, 0.0138) | 0.23 |
|  |  | 3 | 0.0039 (-0.0047, 0.0125) | 0.38 |
|  |  | 4 | 0.0034 (-0.0064, 0.0132) | 0.50 |
| AgeAccelGrim2 | 4.15 | 1 | 0.0169 (0.0085, 0.0254) | <0.01 |
|  |  | 2 | 0.0161 (0.0075, 0.0247) | <0.01 |
|  |  | 3 | 0.0132 (0.0036, 0.0228) | <0.01 |
|  |  | 4 | 0.0155 (0.0041, 0.0270) | <0.01 |

Abbreviations: SD, standard deviation; IEAA, intrinsic epigenetic age acceleration; EEAA, extrinsic epigenetic age acceleration; AgeAccelPheno, phenotypic age acceleration; DunedinPACE, Dunedin Pace of Aging calculated from the Epigenome; AgeAccelGrim2, accelerated DNA methylation GrimAge version 2. Model 1 adjusted for age and time between blood draw and MRI assessment. Model 2 adjusted for age, time interval between blood draw and MRI assessment, race, and ethnicity. Model 3 adjusted for age, time interval between blood draw and MRI assessment, race, ethnicity, hormone trial arm, education, and smoking status. Model 4 adjusted for age, time interval between blood draw and MRI assessment, race, ethnicity, hormone trial arm, education, smoking status, physical activity, body mass index, diabetes, cardiovascular disease, non-melanoma cancer, and blood cell composition. For IEAA or EEAA, blood cell composition was not included in the fully adjusted model (see text).

**Figure S1**. Scatter plots of the association of SPARE- BAA with each of the five epigenetic clocks.

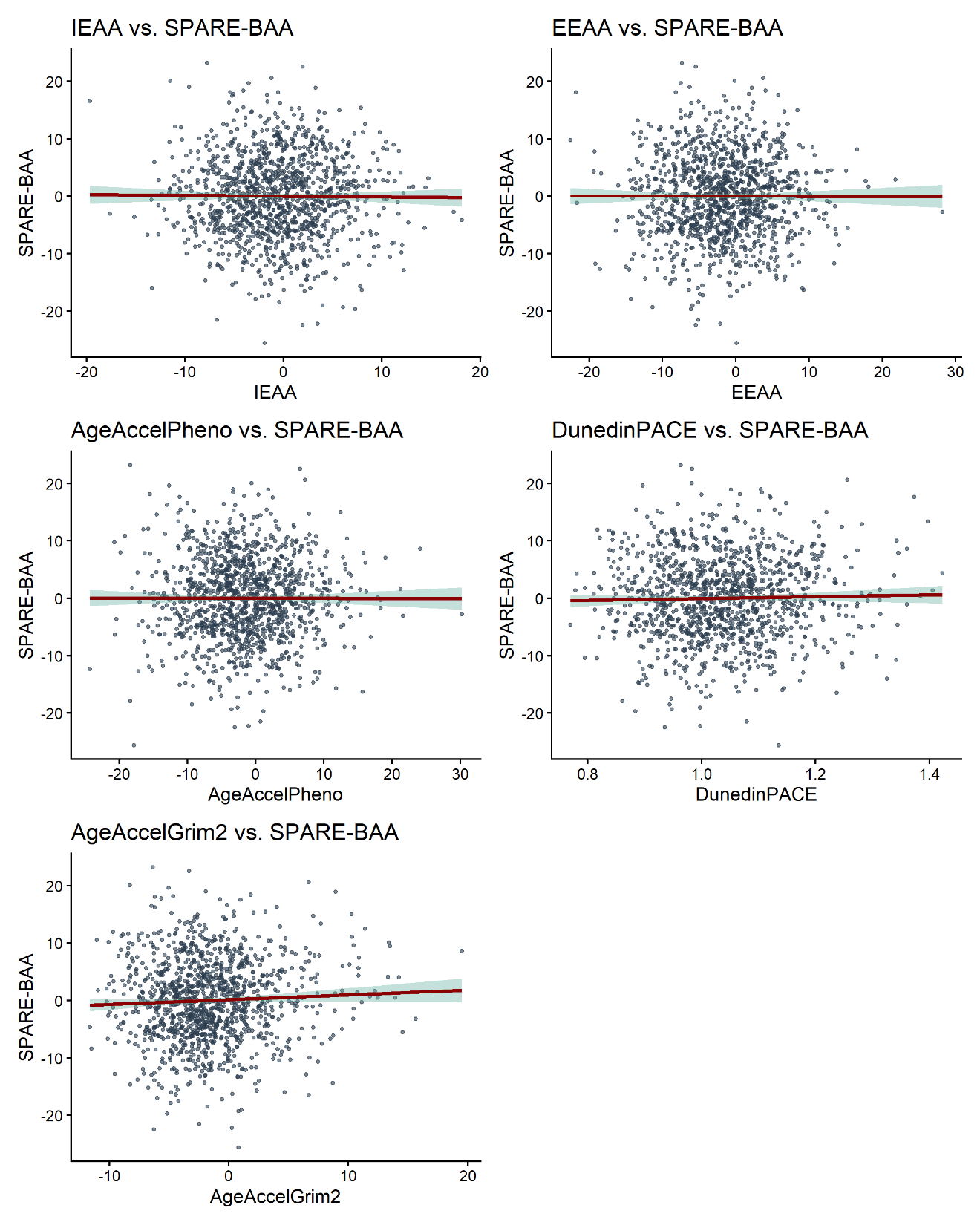

**Figure S2**. Scatter plots of the association of ln(AD-PS + 1) with each of the five epigenetic clocks.

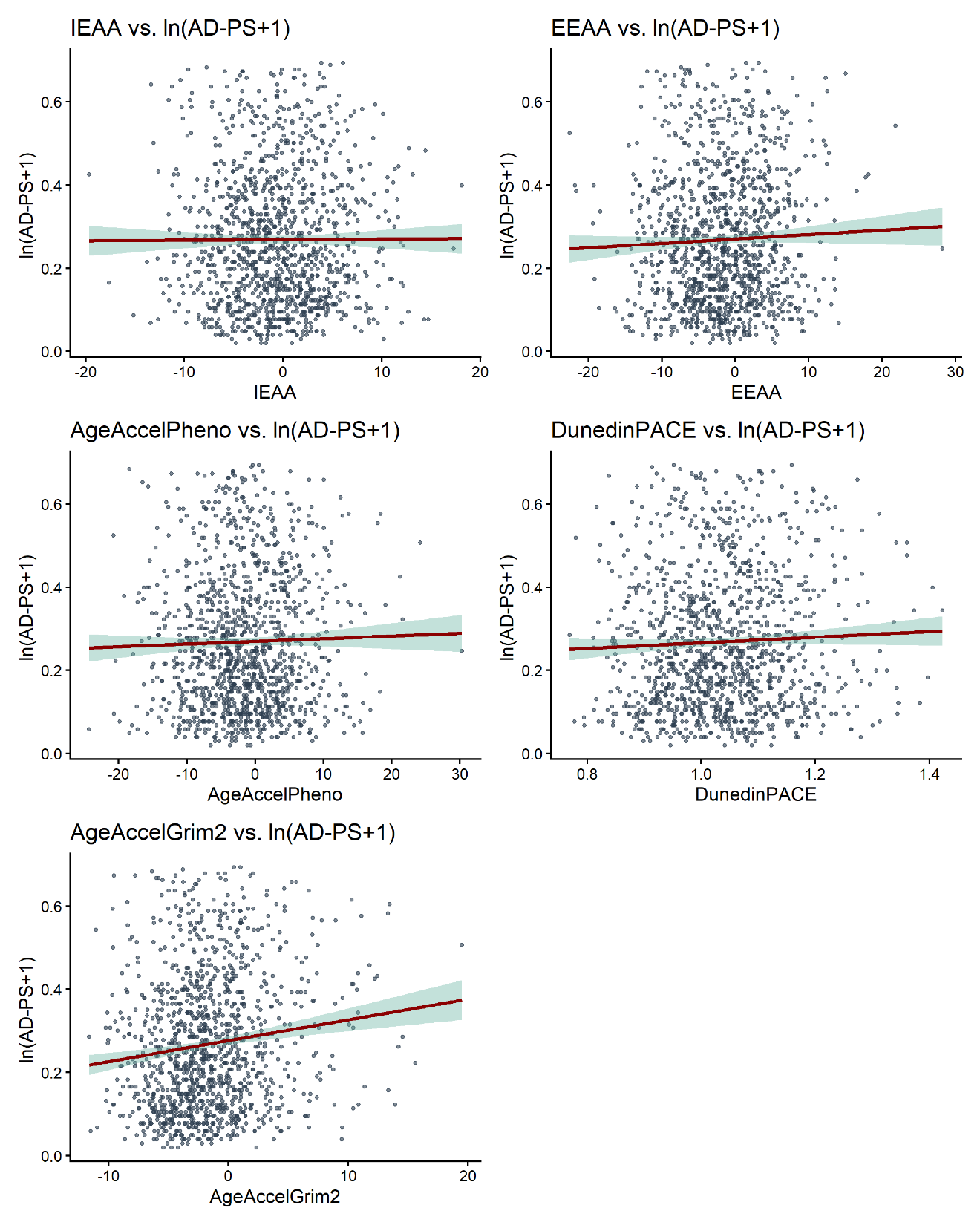

**Figure S3**. Associations of the five epigenetic aging clocks with ln(AD-PS + 1), excluding 46 women who were diagnosed with MCI or AD prior to the MRI scan from linear regression models adjusting for chronological age, hormone therapy trial arm, education, smoking status, race, and ethnicity, physical activity, BMI, diabetes, cardiovascular disease, cancer, and blood cell composition (models for IEAA and EEAA did not include blood cell composition).

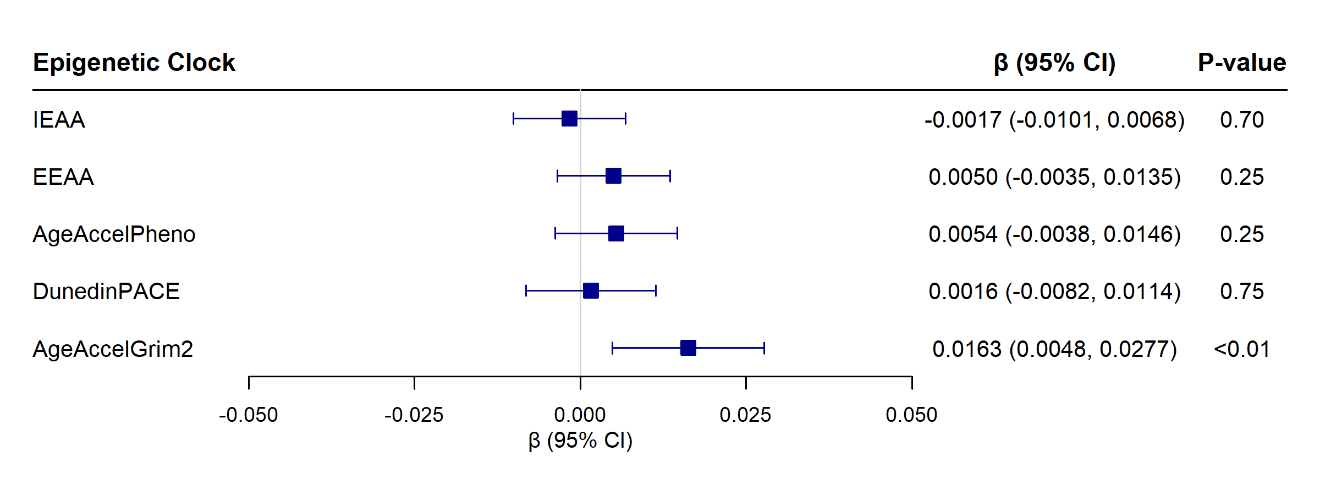
